# Computational Evaluation of a Turbulence-like Electrical Activity Hypothesis in Atrial Fibrillation: Substrate Remodeling, Critical Wavelength Transition, and Multi-wavelet Maintenance

**DOI:** 10.64898/2026.08.08.26360016

**Authors:** Xin Chu, Qing Qiao, Jinpeng Xu, Xiaojun Wang, Mengmeng Li, Chenxi Jiang, Ribo Tang, Tong Liu, Xin Zhao, Hong Ye, Ziwei Xu, Kangning Han, Ping Guo, Biao Fu, Deyong Long

**Author notes:** Corresponding authors: Xin Chu,; Deyong Long.

## Abstract

**BACKGROUND:** Atrial fibrillation (AF) remains difficult to explain using a single focal-driver or rotor-centered mechanism across disease stages. We tested whether progressive atrial substrate remodeling can drive a critical transition toward turbulence-like, decentralized multi-wavelet electrical activity.

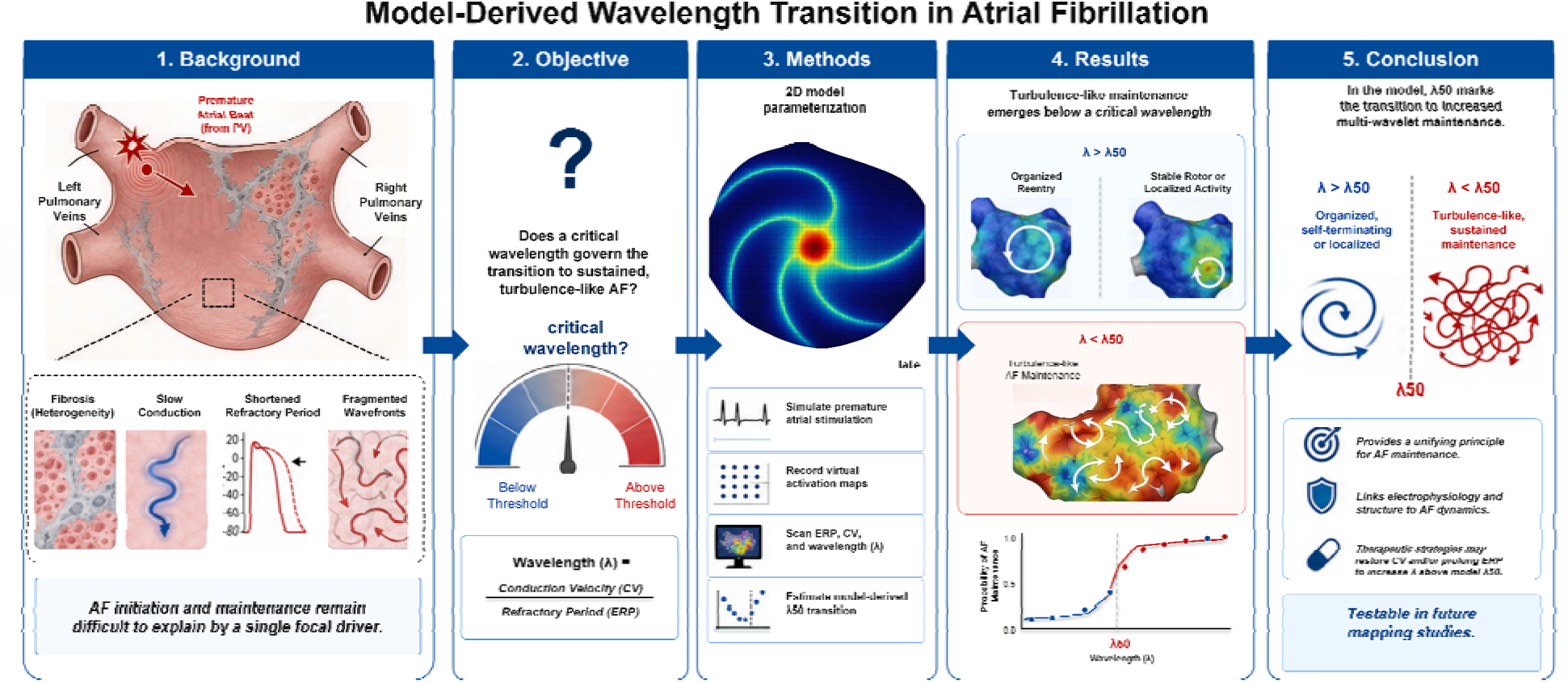

**METHODS:** We constructed a controlled two-dimensional atrial reaction-diffusion model with six graded substrate-remodeling stages. We evaluated effective wavelength, theoretical wavelet capacity, AF inducibility, vulnerable-window dynamics, spatial randomness, temporal memory, spectral dispersion, nonlinear indices, virtual ablation response and ERP-prolongation reverse mechanistic testing.

**RESULTS:** Progressive remodeling shortened effective wavelength from 12.0 to 2.4 cm and increased theoretical wavelet capacity from 0.69 to 17.36. Inducibility rose sigmoidally as wavelength shortened, with a model-derived transition near lambda50=4.5 cm. Advanced substrates showed increased wavebreak, spatial randomness, short-memory dynamics, broad spectral dispersion, positive nonlinear indices and resistance to random local ablation. Culprit atrial premature beats within the vulnerable window efficiently triggered AF, whereas counter-pacing at 20 to 35 ms reduced inducibility from 52% to 11% in stage 2.

**CONCLUSIONS:** In this controlled model, AF initiation and maintenance were linked to substrate-dependent wavelength, wavelet capacity and vulnerable-window triggering. The model-derived transition provides a testable framework for future high-density mapping, patient-specific modeling and device-based studies.

**Clinical Perspective:** *WHAT IS KNOWN?:*

- Pulmonary-vein ectopy, acute autonomic or metabolic triggers and other perturbation sources can initiate paroxysmal or self-limited AF, particularly when they fall into a transient physiological atrial vulnerable window.
- Substrate remodeling with refractory-period shortening, slow conduction and fibrosis is recognized as a key determinant of AF maintenance, but a quantitative wavelength threshold separating trigger-dependent AF from self-maintaining turbulence-like AF has not been established.

*WHAT THE STUDY ADDS:*

- In this controlled two-dimensional model, the inducibility analysis provides a quantitative estimate of an effective transition near 4.5 cm, offering a measurable framework for examining AF maintenance beyond focal-driver or rotor-centered explanations.
- The model links perturbation-source strength, physiological vulnerable-window timing and substrate capacity into a single framework, explaining how apparently physiological AF initiation can become pathological sustained AF when wavelength shortens and wavelet capacity increases.
- A virtual counter-pacing experiment shows that time-locked stimulation after a culprit atrial premature beat can pre-empt local excitability, close the vulnerable window and reduce AF inducibility, suggesting a testable trigger-interception strategy.

## Introduction

Atrial fibrillation (AF) is the most common sustained arrhythmia in clinical practice and is an important cardiovascular phenotype associated with increased risks of stroke, heart failure and death. Lippi et al.^1^ showed, using Global Burden of Disease data, that AF incidence and prevalence have continued to rise over the past two decades and are expected to increase further over the next 30 years. A national cross-sectional study by Shi et al.^2^ reported an age-standardized AF prevalence of approximately 1.6% among Chinese adults, with clear variation by age, sex and region. Lane et al.^3^ noted that AF affects approximately 37.6 million people and will be detected more frequently with the expansion of screening and wearable technologies. Contemporary guidelines have shifted AF care from rhythm or rate control alone to integrated management that includes stroke prevention, risk-factor modification, symptom control and early rhythm intervention.^4,5^ Chinese recommendations have also provided a framework for standardized AF management in China.^6^ Nevertheless, persistent and long-standing persistent AF remain characterized by high recurrence after cardioversion, unstable ablation targets and heterogeneous therapeutic response, suggesting that their maintenance cannot be explained by a single electrophysiological abnormality. Work by Brown et al.^7^ on metabolic, structural, contractile and electrical remodeling also supports the view that AF progression involves multilevel coupling.

Mechanistic theories of AF have evolved from re-entry to multi-wavelet re-entry, rotor-based explanations and nonlinear dynamical frameworks. Studies by Mines,8 Moe and colleagues,9,10 Cox et al.,11 Wang et al.,12 Davidenko et al.13 and Gray et al.14 collectively indicate that AF maintenance cannot be fully explained by a single trigger point, but instead depends on tissue size, effective refractory period (ERP), conduction velocity (CV), spatial heterogeneity and wavefront interactions. Existing theories explain parts of re-entry, rotor or multi-wavelet maintenance, but a unified framework is still needed to quantify wavelength shortening, wavebreak proliferation, spatial decentralization, short-memory dynamics and poor response to local ablation. From a nonlinear-science perspective, this analogy is more than a descriptive metaphor: turbulence theory describes how systems driven far from equilibrium can generate irregular, self-organized and scale-coupled patterns through local interactions, boundary constraints and continuous energy or excitation input. In atrial tissue, heterogeneous conduction velocity, spatially variable refractory recovery and changing re-entry pathways provide a biological substrate in which electrical activation can behave as a complex excitable-medium process rather than as a single deterministic circuit. In this sense, turbulence-like electrical activity should be interpreted as a multiscale nonlinear state linking tissue-scale structure, wavefront fragmentation and cellular recovery dynamics. The turbulence-like electrical activity hypothesis proposed by Chu et al.15 states that, when fibrosis, slow conduction, refractory-period shortening and repolarization dispersion accumulate beyond a critical level, atrial electrical activity may shift from relatively ordered propagation or local driving to a decentralized dynamical state maintained by numerous short-lived wavefronts that are continuously generated, collide, annihilate and regenerate. This state should not be equated with superficial electrogram irregularity. It should be supported by effective-wavelength shortening, increased theoretical wavelet capacity, wavebreak proliferation, randomized spatial distribution, rapid decay of temporal correlation, spectral dispersion, increased nonlinear chaotic components and reduced response to local ablation. On this basis, the present study constructed a stage 0 to stage 5 graded substrate-remodeling model and tested the evidence chain from substrate remodeling to critical wavelength shortening, wavebreak proliferation and decentralized multi-wavelet maintenance across induction threshold, spatial organization, temporal memory, spectral behavior, nonlinear dynamics and intervention response. The overall computational evaluation workflow is summarized in Figure 1.

**Figure 1.**
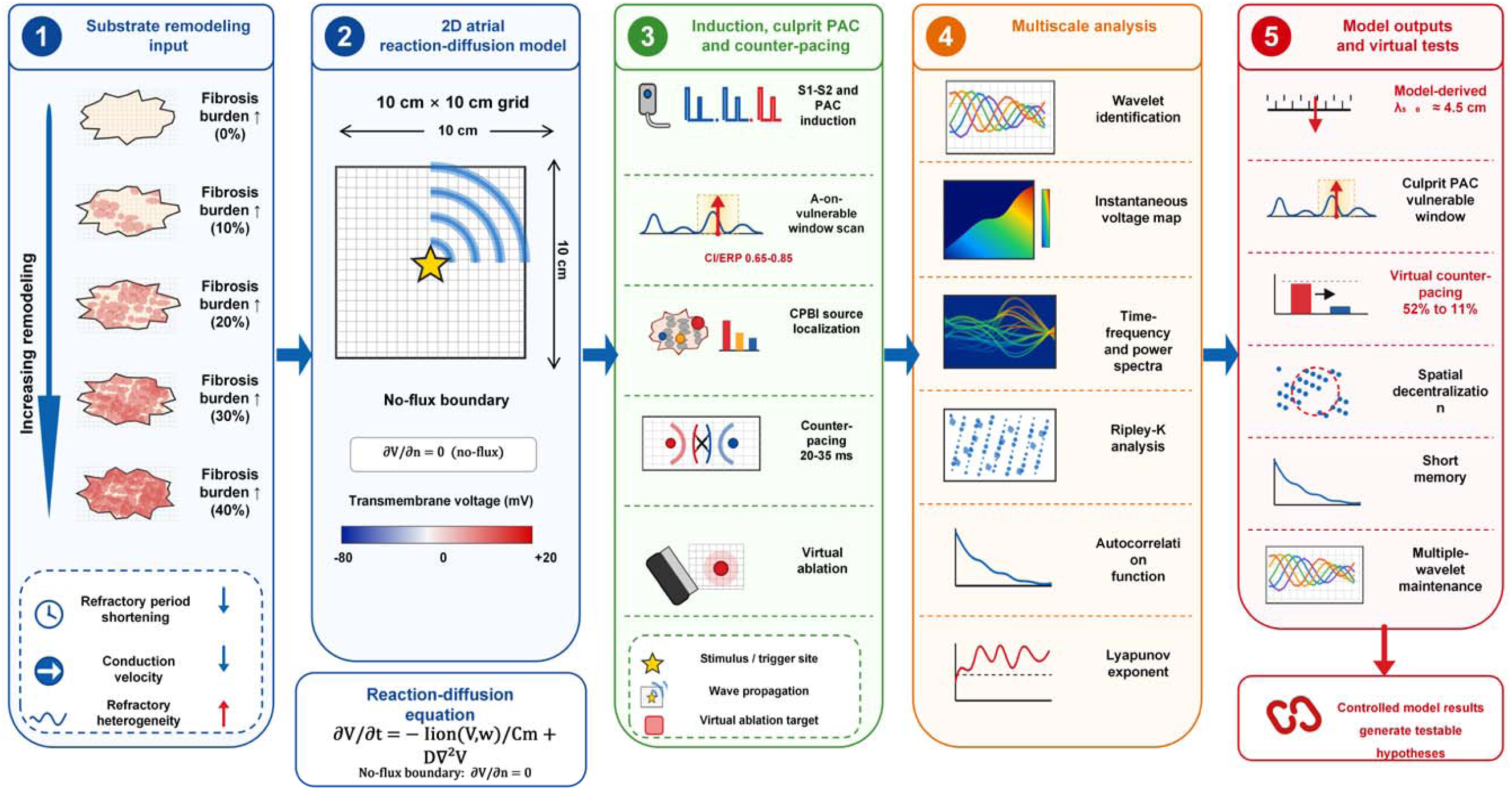
Overall workflow of the computational evaluation study.

## Methods

### Study Design and Hypothesis-Testing Pathway

This was a hypothesis-testing computational simulation study using a two-dimensional atrial excitable-tissue model and the spatiotemporal electrophysiological data generated by that model. The aim was to test the AF turbulence-like electrical activity hypothesis within a controlled parameter space. The overall pathway included four layers: first, construction of graded remodeling models from normal control to advanced AF substrate; second, identification of the induction threshold for a turbulence-like state through single premature atrial beat stimulation and wavelength scanning; third, assessment of whether the induced state met decentralized multi-wavelet criteria using spatial, temporal, spectral and nonlinear-dynamical indices; and fourth, virtual ablation and counter-pacing experiments to determine whether the state depended on a single driver and whether it could be prevented at the triggering stage. Key substrate parameters are shown in Table 1 and Figure 1.

**Table 1.** Key parameter configuration of the six-stage atrial substrate-remodelling model.

| Stage | Simulated state | pfib (%) | rERP (%) | rCV (%) | ERP (ms) | CV (cm/s) | $\lambda$ (cm) | Ncritical |
| --- | --- | --- | --- | --- | --- | --- | --- | --- |
| Stage 0 | Normal control | 0 | 0 | 0 | 200 | 60 | 12 | 0.69 |
| Stage 1 | Early electrical remodelling | 0 | 20 | 0 | 160 | 60 | 9.6 | 1.08 |
| Stage 2 | Early composite remodelling | 10 | 30 | 0 | 140 | 60 | 8.4 | 1.42 |
| Stage 3 | Early persistent AF | 20 | 40 | 20 | 120 | 48 | 5.76 | 3.01 |
| Stage 4 | Long-standing persistent AF | 30 | 50 | 40 | 100 | 36 | 3.6 | 7.72 |
| Stage 5 | Advanced AF substrate | 40 | 50 | 60 | 100 | 24 | 2.4 | 17.36 |

### Two-Dimensional Atrial Reaction-Diffusion Model

The two-variable system was selected as a minimal excitable-medium model: it retains threshold excitation, recovery and refractoriness while allowing transparent, large-scale interrogation of wavelength-dependent wavebreak dynamics. It was not intended to reproduce ion-channel-specific kinetics or predict drug effects.

### Parameterization of Substrate Remodeling

Substrate remodeling was controlled by fibrosis-mask burden (pfib), refractory-period shortening (rERP), conduction-velocity reduction (rCV), and phenomenological local recovery-time heterogeneity. For standardization, the randomized spatial distribution was used as the baseline configuration to isolate wavelength-dependent effects and reduce confounding by fibrosis geometry; patchy and confluent configurations were not evaluated in this study. The fibrosis mask served as a randomized structural conduction-block surrogate, rather than a reconstruction of replacement, infiltrative, endomysial, patchy, or confluent human fibrosis. The graded model included stages 0 to 5: stage 0 represented normal control, stage 1 early electrical remodeling, stage 2 early composite remodeling, stage 3 early persistent AF, stage 4 long-standing persistent AF and stage 5 advanced remodeling substrate. ERP, CV, effective wavelength lambda and theoretical wavelet capacity Ncritical are listed in Table 1. The ranges of ERP, CV and fibrosis burden were selected to provide clinically informed, literature-based context for this controlled scan (see Supplementary Table S1 for source ranges and modeling roles); they were not fitted to individual patients.

### Stimulation Protocol, Wavelength Scanning and Wavelet Identification

The model used an S1-S2-like perturbation protocol and single premature atrial beat induction. Effective wavelength λ was estimated as the product of ERP and CV. Theoretical wavelet capacity N_critical_ was estimated as the ratio of tissue area to λ^2^. To quantify the transition threshold, λ was set to 8.0, 7.0, 6.0, 5.5, 5.0, 4.5, 4.0, 3.5, 3.0, 2.5 and 2.0 cm, with 50 repeated inductions at each level. Successful induction was defined as persistence of at least three wavelets during the 10 s observation window and at least two independent wavelets remaining at the end of observation. Wavefront identification used nodes with voltage above –50 mV as candidate activated regions, combined with a spatial-gradient threshold of 5 mV/mm to define wavefront boundaries. After eight-connected-component analysis, connected clusters with an area greater than 25 nodes were defined as independent wavelets.

### Physiological Atrial Vulnerable Period, Perturbation-Source Premature Beats and Virtual Experiments

To incorporate occasional AF in an apparently normal atrium into the same dynamical framework, the model introduced a physiological vulnerable window into the S1-S^2^ protocol. Ti denoted the local recovery time of the i-th atrial grid unit after baseline activation and followed a recovery-time distribution with mean ERP and standard deviation sigmarec. When the S^2^ coupling interval CI arrived, tissue was divided into three regions: refractory region PR(CI)=1-FT(CI), slow-conduction region PS(CI)=FT(CI)-FT(CI-Delta s) and fully excitable region PE(CI)=FT(CI-Delta s), where Delta s represented the slow-conduction band during which sodium-channel availability and local conduction safety factor had not fully recovered. The physiological vulnerable-window index was therefore defined as VWI(CI)=PR(CI)*PS(CI)*PE(CI). VWI increases only when refractory, slow-conduction and excitable regions coexist, and therefore quantifies the transient mixed state in which tissue is partly excitable, partly slowly conducting and partly refractory.

Perturbation-source strength was defined as Dsource=APAC*χPV*χauto, where APAC represented premature-beat stimulus amplitude or local recruitment capacity, χPV represented the anatomical triggering weight of pulmonary-vein-origin beats, and χauto represented amplification of excitability and repolarization dispersion by acute factors such as sympathetic activation, alcohol, hypoxia, hyperthyroidism or electrolyte disturbance. VWI was scanned across CI under normal physiological recovery, acute sympathetic/alcohol/hyperthyroidism-like perturbation and pathological substrate remodeling. AF triggering was expressed as Dsource*VWI(CI)>θinit. Whether AF was sustained depended on substrate capacity, namely whether Ncritical=A/λ2 exceeded the threshold required for multi-wavelet maintenance. This configuration allowed physiological self-limited AF and pathological sustained AF to be explained within the same perturbation-source, vulnerable-window and substrate-capacity framework.

### Culprit Premature Atrial Beats and Counter-Pacing Within the Atrial Vulnerable Window

To convert the physiological vulnerable window from an abstract index into a testable triggering event, S1-S2 scanning was further performed in stage 2 and stage 3 substrate models. S2 was defined as a premature atrial beat, with CI/ERP set from 0.50 to 0.95 in steps of 0.05. Each CI/ERP level was repeated 100 times. Because the surface ECG T wave mainly represents ventricular repolarization, this phenomenon was not described as a true P-on-T event. Instead, we used an atrial-vulnerable-window analogy, in which a premature atrial beat falls within a window created by heterogeneous atrial repolarization recovery.

The culprit premature beat index was defined as CPBI(CI)=VWInorm(CI)*(APAC/Acritical)*χPV*χauto. VWInorm(CI) represented the normalized physiological vulnerable-window index. APAC/Acritical represented the ratio of premature-beat recruitment area to the minimal wavefront area needed to form re-entry. χPV represented the anatomical triggering weight of the pulmonary-vein antrum, and χauto represented amplification of excitability and repolarization dispersion by sympathetic activation, alcohol, hypoxia or hyperthyroidism-like states. Premature-beat origins were assigned to the pulmonary-vein antrum, left atrial posterior wall or right atrium to compare CPBI across spatial sources. The counter-pacing experiment was triggered after identification of a culprit beat. Counter-pacing pulses were delivered 10-50 ms after the culprit beat in 5 ms increments, with 25 ms as the primary analysis delay. The counter-pacing region was set within 2 cm of the premature-beat origin, with voltage 2.0 V and pulse width 1.5 ms. The primary endpoint was AF induction success within the virtual observation window. Preventive efficacy was defined as (control AF inductions – AF inductions after counter-pacing)/control AF inductions. Between-group inducibility was compared using Fisher exact tests.

### Multiscale Criteria for Turbulence-like Features

The turbulence-like state was evaluated across five dimensions. The spatial dimension used instantaneous voltage maps, wavelet-centroid distribution and Ripley-K functions to test whether wavelets followed complete spatial randomness. The temporal dimension used wavelet-count time series, autocorrelation and lifetime distribution to test for short memory and memoryless annihilation. The spectral dimension used virtual-electrode continuous wavelet spectra and Welch power spectra to calculate dominant frequency, spectral width and organization index. The nonlinear-dynamical dimension used correlation dimension, maximum Lyapunov exponent and surrogate-data testing to distinguish low-dimensional deterministic chaos from linear stochastic noise. The intervention-response dimension used virtual ablation and counter-pacing to compare intervenability during the maintenance and triggering phases, respectively.

### Model Quality Control and Reproducibility

To reduce dependence on a single random fibrosis realization, experiments involving randomized structures were repeated across independent realizations, and mean, standard deviation, standard error and 95% confidence intervals were reported at the summary level. Spatial discretisation used a fixed grid scale, and temporal integration used a fixed time step. All stages were compared under the same tissue area, boundary conditions and wavelet-identification rules, so that inter-stage differences primarily reflected substrate-parameter changes rather than numerical settings. In the wavelength-scanning experiment, each wavelength level was repeated 50 times and induction success was calculated using identical criteria. For analysis of the stage 5 turbulence-like state, spatial, temporal, spectral, chaotic and intervention-response indices were used for cross-checking. A state showing only a broad spectrum without spatial randomness, or only multiple wavelets with long-term periodic autocorrelation, was not classified as turbulence-like by itself.

### Statistical Analysis

All statistical analyses were performed using R version 4.5.2. The analyses used trial-level and summary outputs generated from the virtual simulation workflow. No patient-level data were analyzed. All tests were two-sided unless otherwise specified, and P<0.05 was considered statistically significant.

Continuous simulation outputs were summarized as mean, standard deviation, standard error and 95% confidence interval when applicable. Proportional endpoints, including AF inducibility, wavebreak incidence, termination rate and persistence rate, were reported as event counts and percentages, with Wilson 95% confidence intervals for binomial proportions. Effective wavelength was calculated as ERP multiplied by CV, and theoretical wavelet capacity was calculated as Ncritical = A/lambda2. The relationship between effective wavelength and AF inducibility was analyzed using a binomial logistic/sigmoidal model, with the number of induced and non-induced trials as the binomial response. lambda50 was defined as the wavelength corresponding to a predicted 50% induction probability, with the observed grid-level transition used as a sensitivity check.

For culprit premature atrial beat and counter-pacing experiments within the atrial vulnerable window, CI/ERP scanning was repeated 100 times at each CI/ERP level in stages 2 and 3. AF inducibility after culprit premature atrial beats alone and after 25-ms counter-pacing was compared using Fisher exact tests. Absolute risk reduction and relative preventive efficacy were calculated from the trial-level induction counts. Counter-pacing delay was analyzed from 10 to 50 ms, and the prespecified optimal response window was 20-35 ms. CPBI values were summarized at the trial level for each anatomical origin and compared across origins using one-way ANOVA, with Kruskal-Wallis testing as a distribution-robust sensitivity analysis. Pairwise origin comparisons were adjusted using the Benjamini-Hochberg false-discovery-rate procedure.

Spatial organization was assessed using wavelet-centroid point-process analysis, Ripley-K functions with Monte Carlo confidence envelopes and a wavelet-location chi-square test. Temporal memory was quantified from the wavelet-count autocorrelation function, and wavelet-lifetime distributions were evaluated using the Kolmogorov-Smirnov test against the exponential distribution. Spectral analyses used virtual-electrode time-frequency spectra and Welch power spectra to derive dominant frequency, spectral width and organization index. Nonlinear dynamics were assessed using correlation dimension, maximum Lyapunov exponent and surrogate-data testing. Virtual-ablation endpoints were analyzed as proportions or time constants according to endpoint type. No patient-level data were used, and no missing-data imputation was performed.

## Results

### Graded Remodeling Shifted Tissue From Low Wavelet Capacity to Multi-Wavelet Maintenance

The stage-wise parameter framework is summarized in Table 1 and Figure 2. In stage 0, ERP was 200 ms, CV was 60 cm/s, effective wavelength was 12.0 cm and Ncritical was only 0.69. With increasing remodeling, wavelength shortened to 5.76 cm in stage 3, while Ncritical increased to 3.01. In stages 4 and 5, wavelength decreased to 3.6 cm and 2.4 cm, respectively, and Ncritical increased to 7.72 and 17.36. Raw summary data showed that stable wavelet count increased progressively from 0.1 in stage 0 to 7.2 in stage 5, whereas wavebreak incidence increased from 18.5% in stage 2 to 52.3% in stage 3, 78.6% in stage 4 and 91.2% in stage 5 (Table 2 and Figure 3).

**Figure 2.**
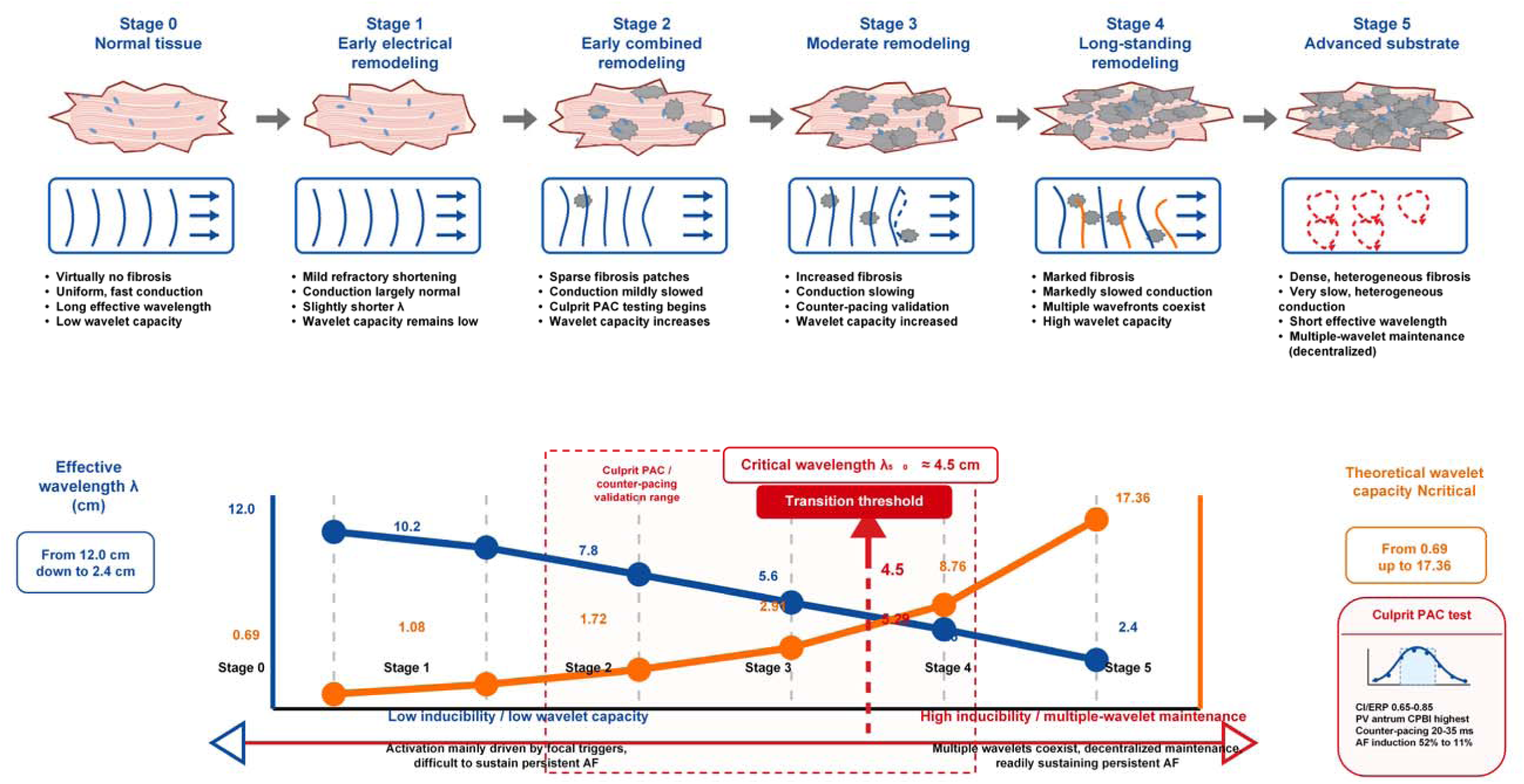
Graded substrate-remodelling parameter gradient and critical mass.

**Figure 3.**
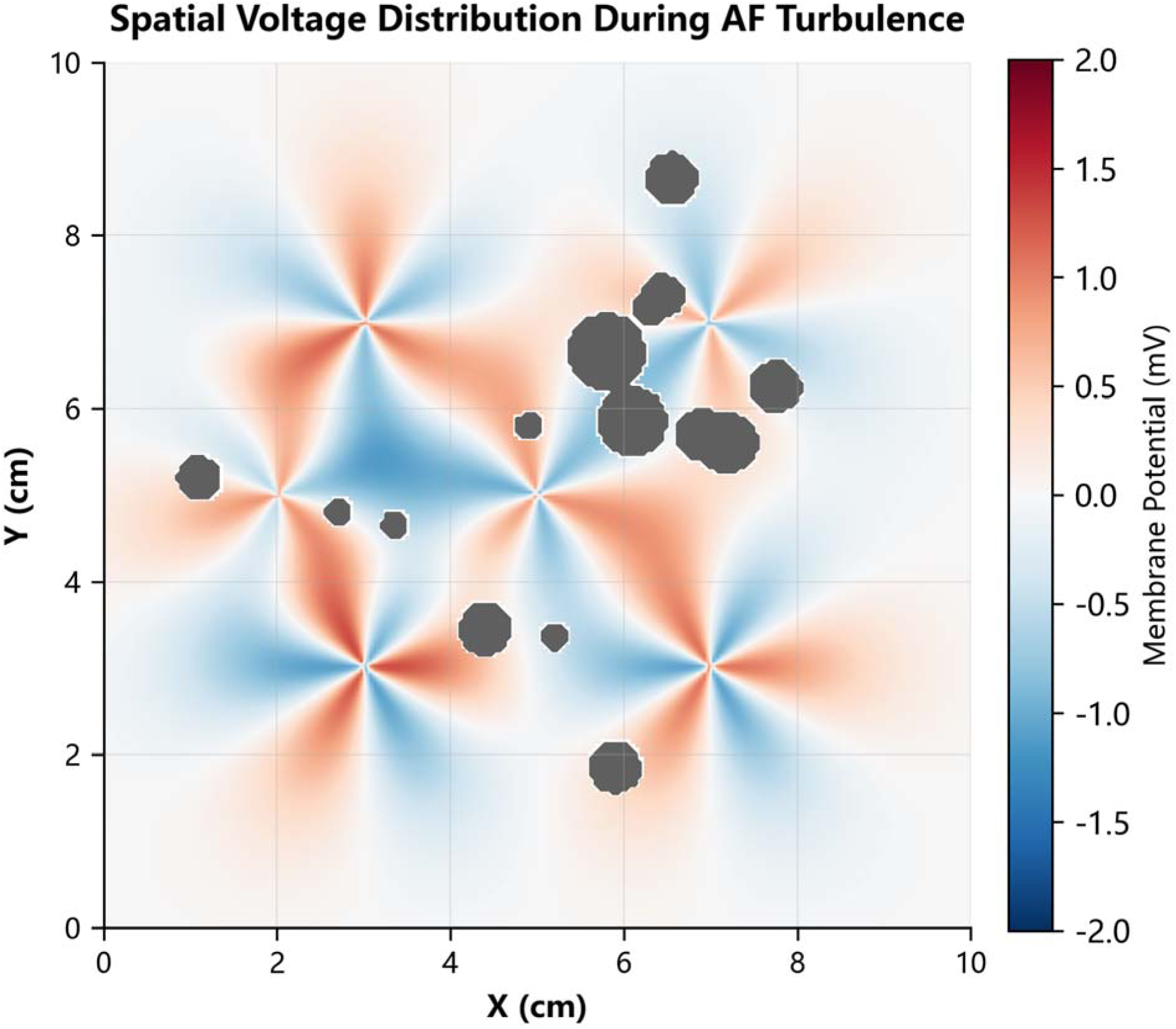
Instantaneous voltage distribution in the stage 5 advanced substrate. Multiple independent wavefronts fragmented, detoured and regenerated around fibrotic boundaries, forming a decentralised multi-wavelet state.

**Table 2.** Wavefront fragmentation and wavelet-maintenance indices under graded remodelling.

| Stage | $\lambda$ (cm) | Ncritical | Stable wavelet count (95% CI) | Wavebreak incidence (95% CI) |
| --- | --- | --- | --- | --- |
| Stage 0 | 12 | 0.69 | 0.1(0.06-0.14) | — |
| Stage 1 | 9.6 | 1.08 | 0.8(0.66-0.94) | — |
| Stage 2 | 8.4 | 1.42 | 1.5(1.28-1.72) | 18.5%(18.0-19.0) |
| Stage 3 | 5.76 | 3.01 | 3.2(2.89-3.51) | 52.3%(51.7-52.9) |
| Stage 4 | 3.6 | 7.72 | 5.8(5.44-6.16) | 78.6%(78.2-79.0) |
| Stage 5 | 2.4 | 17.36 | 7.2(6.78-7.62) | 91.2%(90.9-91.5) |

### Critical Wavelength Showed a Phase-Transition-like Induction Threshold

Wavelength scanning showed a nonlinear sigmoidal increase in induction success as λ shortened (Table 3 and Figure 4). When λ was at least 5.5 cm, induction success remained low at 2-8%. When λ decreased to 5.0 cm, induction success increased to 26%. At λ=4.5 cm, success rose sharply to 58%. Once λ was 4.0 cm or less, success continued to increase and reached 100% at λ=2.0 cm. Based on raw summary statistics, λ50 was approximately 4.5 cm, with a 95% confidence interval of 4.47-4.53 cm.

**Figure 4.**
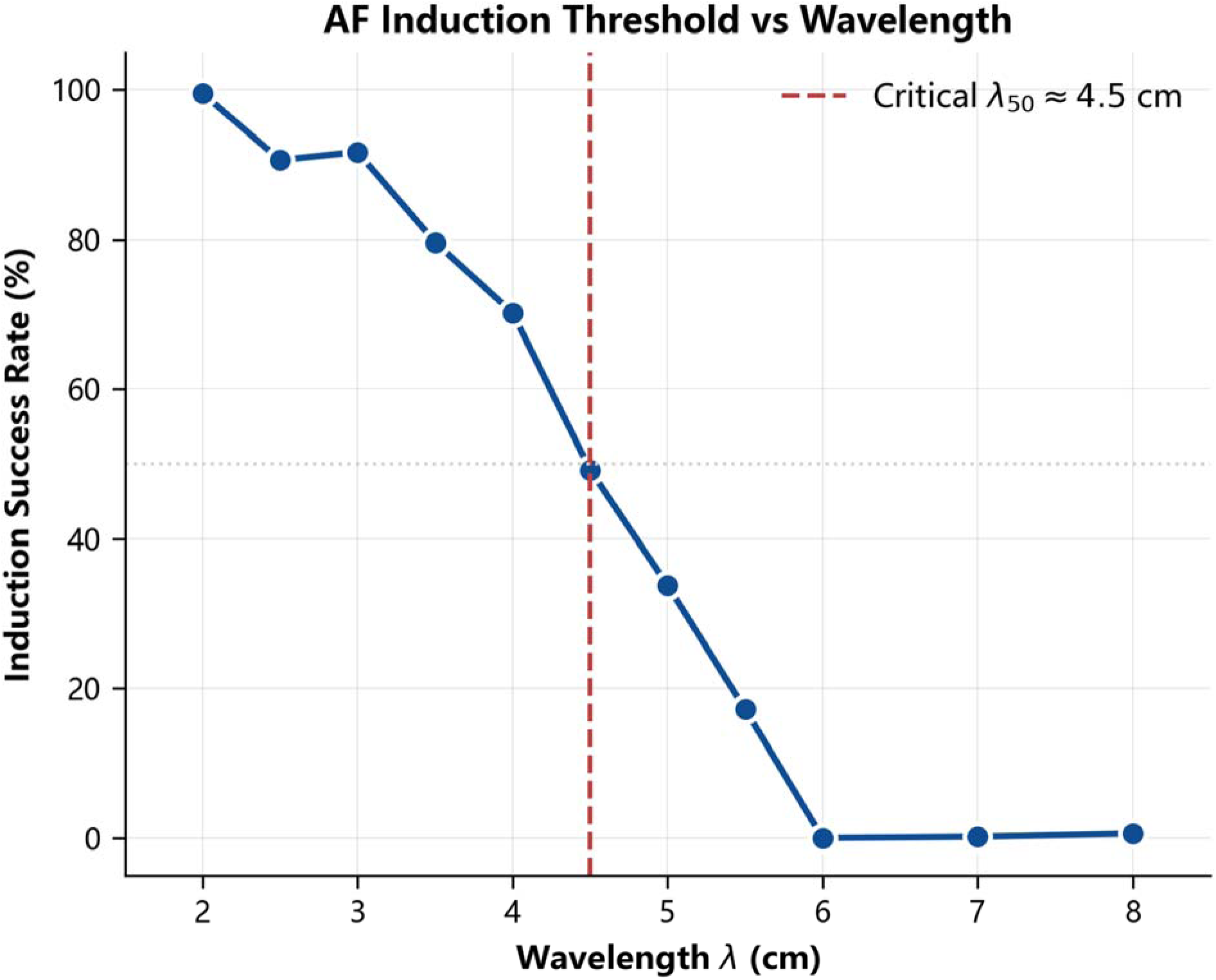
Sigmoidal relationship between turbulence-like state inducibility and effective wavelength. The estimated λ_50_was approximately 4.5 cm, marking the transition from low to high inducibility.

**Table 3.** Induction success rate of the turbulence-like state under different effective wavelengths. Induction proportions are shown with 95% Wilson confidence intervals. The wavelength effect was tested using binomial logistic regression of AF inductions/trials against effective wavelength (P = 6.99e-34***). *P<0.05, **P<0.01 and ***P<0.001.

| $\lambda$ (cm) | Trials | Successes | Induction success (%) | 95% CI |
| --- | --- | --- | --- | --- |
| 8 | 50 | 1 | 2 | 0-5.9 |
| 7 | 50 | 4 | 8 | 0.5-15.5 |
| 6 | 50 | 2 | 4 | 0-9.4 |
| 5.5 | 50 | 3 | 6 | 0-12.6 |
| 5 | 50 | 13 | 26 | 13.8-38.2 |
| 4.5 | 50 | 29 | 58 | 44.3-71.7 |
| 4 | 50 | 35 | 70 | 57.3-82.7 |
| 3.5 | 50 | 42 | 84 | 73.8-94.2 |
| 3 | 50 | 44 | 88 | 79-97 |
| 2.5 | 50 | 47 | 94 | 87.4-100 |
| 2 | 50 | 50 | 100 | 100-100 |

### Virtual Assessment of the Physiological Vulnerable Window and Perturbation-Source Premature Beats

The virtual experiment showed that a narrow VWI peak can appear during repolarization recovery after a single depolarization in the normal atrium. This interval corresponds to the coexistence of refractory, slow-conduction and fully excitable regions. If a culprit premature beat from the pulmonary veins or another trigger site arrives within this window, the perturbation source can generate transient re-entry or multi-wavelet activity through local one-way block and slow conduction. However, because Ncritical is low in a stage 0-like atrium, most such events are predicted to be self-limited. Acute sympathetic activation, alcohol exposure or hyperthyroidism-like conditions can widen or left-shift the VWI curve by shortening mean ERP, increasing recovery dispersion and increasing perturbation-source gain, thereby explaining why paroxysmal AF may occur in some patients without marked chronic structural remodeling. By contrast, stage 3 to stage 5 pathological substrate mainly increases Ncritical and reduces effective wavelength, making the same perturbation more likely to progress to sustained maintenance (Figure 5).

**Figure 5.**
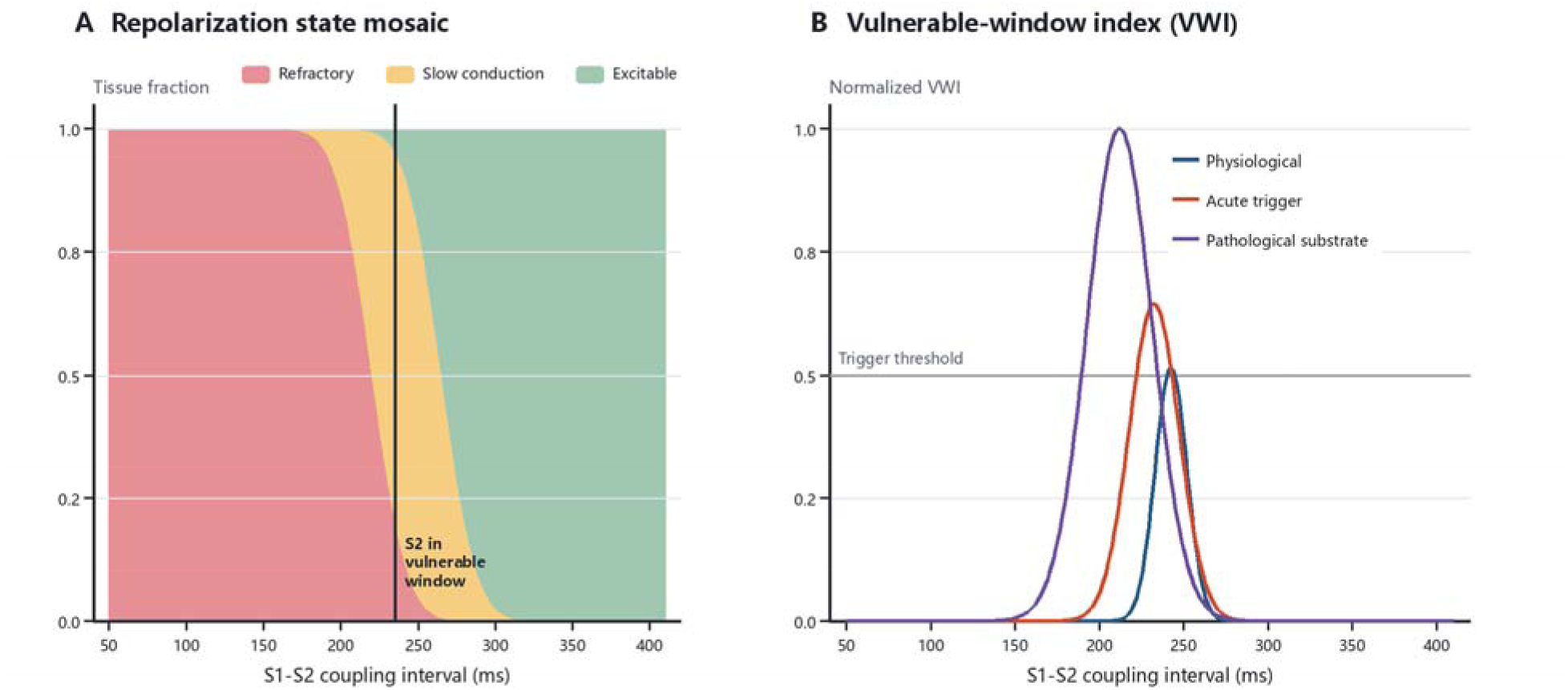
Physiological atrial vulnerable period and vulnerable-window index. (A) After baseline activation, incomplete recovery creates a time window in which refractory, slow-conduction and excitable regions coexist. (B) The normal vulnerable window is narrow; acute triggers widen or left-shift the window, whereas pathological substrate increases the probability of AF maintenance.

### Virtual Assessment of Culprit Premature Atrial Beats and Counter-Pacing Within the Atrial Vulnerable Window

S1-S2 scanning showed clear phase specificity in AF induction by culprit premature atrial beats. In the stage 2 substrate, AF inducibility rose markedly when CI/ERP was 0.65-0.85 and reached 52% at CI/ERP=0.75. At CI/ERP=0.50-0.60 or 0.90-0.95, inducibility was only 4-10%. The stage 3 substrate showed the same pattern but higher vulnerability, with inducibility of 78% at CI/ERP=0.75. CPBI also differed by premature-beat origin. Beats originating from the pulmonary-vein antrum had the highest CPBI (0.83±0.09), compared with the left atrial posterior wall (0.52±0.11) and right atrium (0.37±0.08). These findings indicate that a culprit premature atrial beat is determined not only by prematurity, but also by spatial recruitment capacity and anatomical triggering weight. Local counter-pacing delivered 25 ms after the culprit beat significantly reduced AF inducibility. In stage 2, inducibility fell from 52% to 11%, an absolute reduction of 41%. In stage 3, inducibility fell from 78% to 34%, an absolute reduction of 44%. Fisher exact tests were significant in both models (P<0.001). The preventive effect of counter-pacing was time-window dependent. In stage 2, preventive efficacy was 11.5% and 23.1% at 10 and 15 ms delays, respectively. At 20-35 ms, efficacy increased to 71.2-82.7%, peaking at 82.7% at 25 ms. At 45-50 ms, efficacy decreased to 25.0-13.5% (Figure 6, Figure 7 and Table 4).

**Figure 6.**
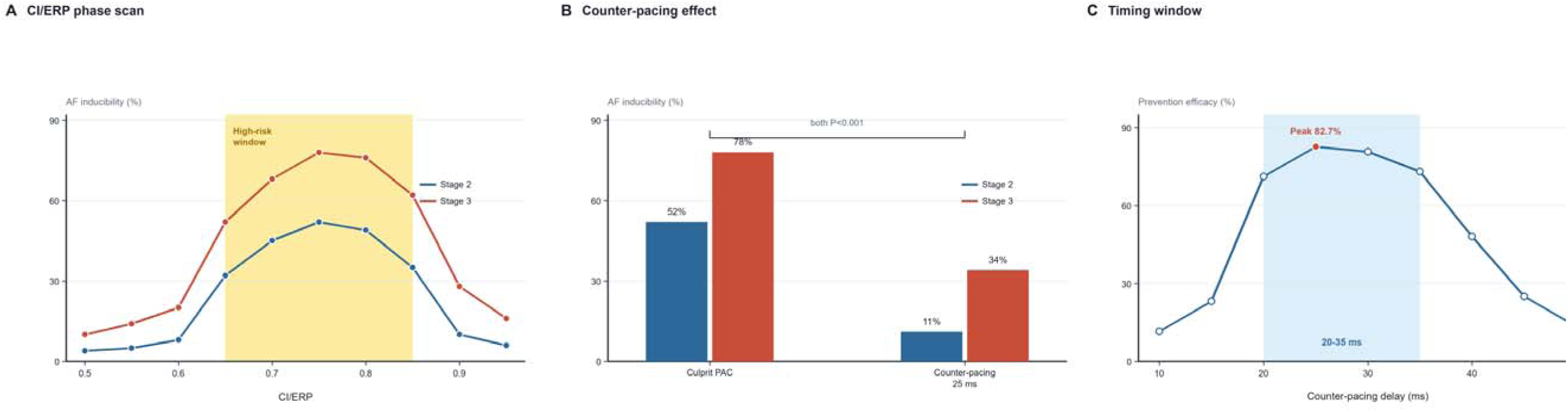
Statistical results for culprit premature atrial beats and counter-pacing within the atrial vulnerable window. (A) CI/ERP scanning identified 0.65-0.85 as the high-inducibility window, with higher overall vulnerability in stage 3 than in stage 2. (B) Counter-pacing at 25 ms significantly reduced AF inducibility in both stage 2 and stage 3 models (Fisher exact test, both P<0.001). (C) The preventive effect was time-window dependent, with the optimal window at 20-35 ms.

**Figure 7.**
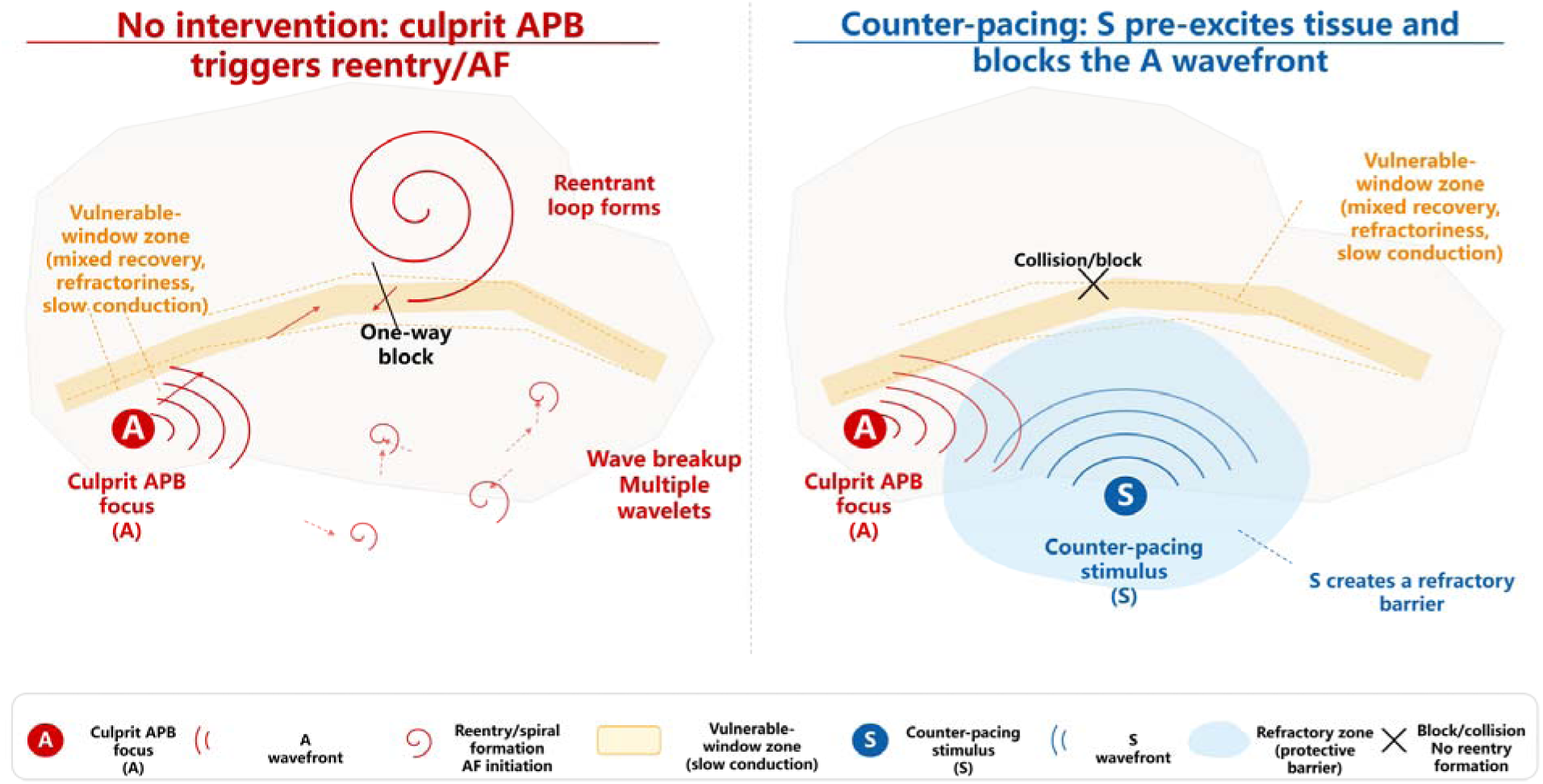
Mechanistic schematic showing how counter-pacing blocks the abnormal wavefront generated by a culprit premature atrial beat. Point A denotes the culprit APB focus and point S denotes the counter-pacing stimulus. S pre-excites surrounding tissue and creates a refractory barrier, thereby blocking or colliding with the abnormal A wavefront before it reaches the critical slow-conduction zone.

**Table 4.** Virtual assessment results for culprit premature atrial beats and counter-pacing within the atrial vulnerable window. P values were calculated using two-sided Fisher exact tests comparing AF inducibility after culprit premature atrial beats alone with AF inducibility after 25-ms counter-pacing within the same substrate stage. Reference rows indicate the corresponding culprit APB-only control. *P<0.05, **P<0.01 and ***P<0.001.

| Model stage/analysis | Condition | Trials | AF inductions | Inducibility or efficacy | P value |
| --- | --- | --- | --- | --- | --- |
| Stage 2 | Culprit APB (CI/ERP=0.75) | 100 | 52 | 52.0% | Reference |
| Stage 2 | Counter-pacing at 25 ms | 100 | 11 | 11.0% | $P = 3.73\text{e-}10^{***}$ |
| Stage 3 | Culprit APB (CI/ERP=0.75) | 100 | 78 | 78.0% | Reference |
| Stage 3 | Counter-pacing at 25 ms | 100 | 34 | 34.0% | $P = 4.59\text{e-}10^{***}$ |
| Stage 2 timing-window analysis | Counter-pacing delay of 25 ms | 100 | 9 | Preventive efficacy 82.7% | Not tested; peak window |

### Spatial Randomness and Short-Memory Dynamics Supported Decentralized Maintenance

Stage-wise wavelet-count statistics showed a mean of 6.45 wavelets in stage 5, with a standard deviation of 2.18 and interquartile range of 5-8. Stage 4 already reached the lower limit for multi-wavelet maintenance, with a mean wavelet count of 4.94 (Table 5). Stage 5 instantaneous voltage maps showed multiple independent wavefronts detouring, fragmenting and regenerating around fibrotic boundaries, without a single fixed centre driving the entire tissue. In the wavelet-centroid spatial point-process analysis, the Ripley-K test gave P=0.42 and the wavelet-location chi-square test gave P=0.38, so complete spatial randomness was not rejected (Table 6 and Figure 8). The corresponding autocorrelation analysis showed rapid short-memory decay in the turbulence-like state (Figure 9). Temporally, the ACF correlation time was 348 ms. Mean wavelet lifetime was 352 ms, and the Kolmogorov-Smirnov test gave P=0.35, supporting an exponential lifetime distribution.

**Figure 8.**
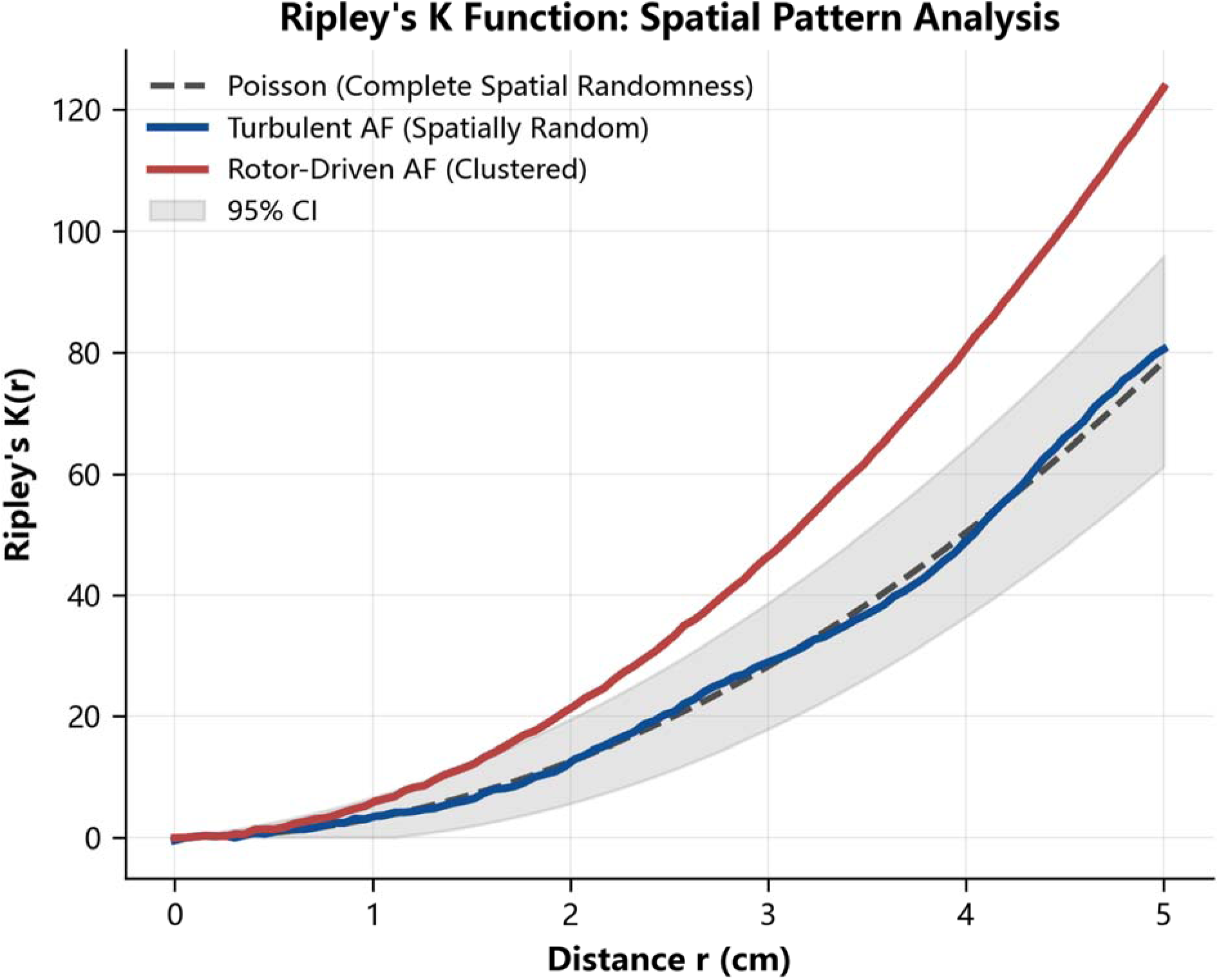
Ripley-K spatial point-process analysis. The turbulence-like state approximated the theoretical Poisson random-process line and remained within the 95% confidence envelope, supporting decentralised spatial wavelet distribution.

**Figure 9.**
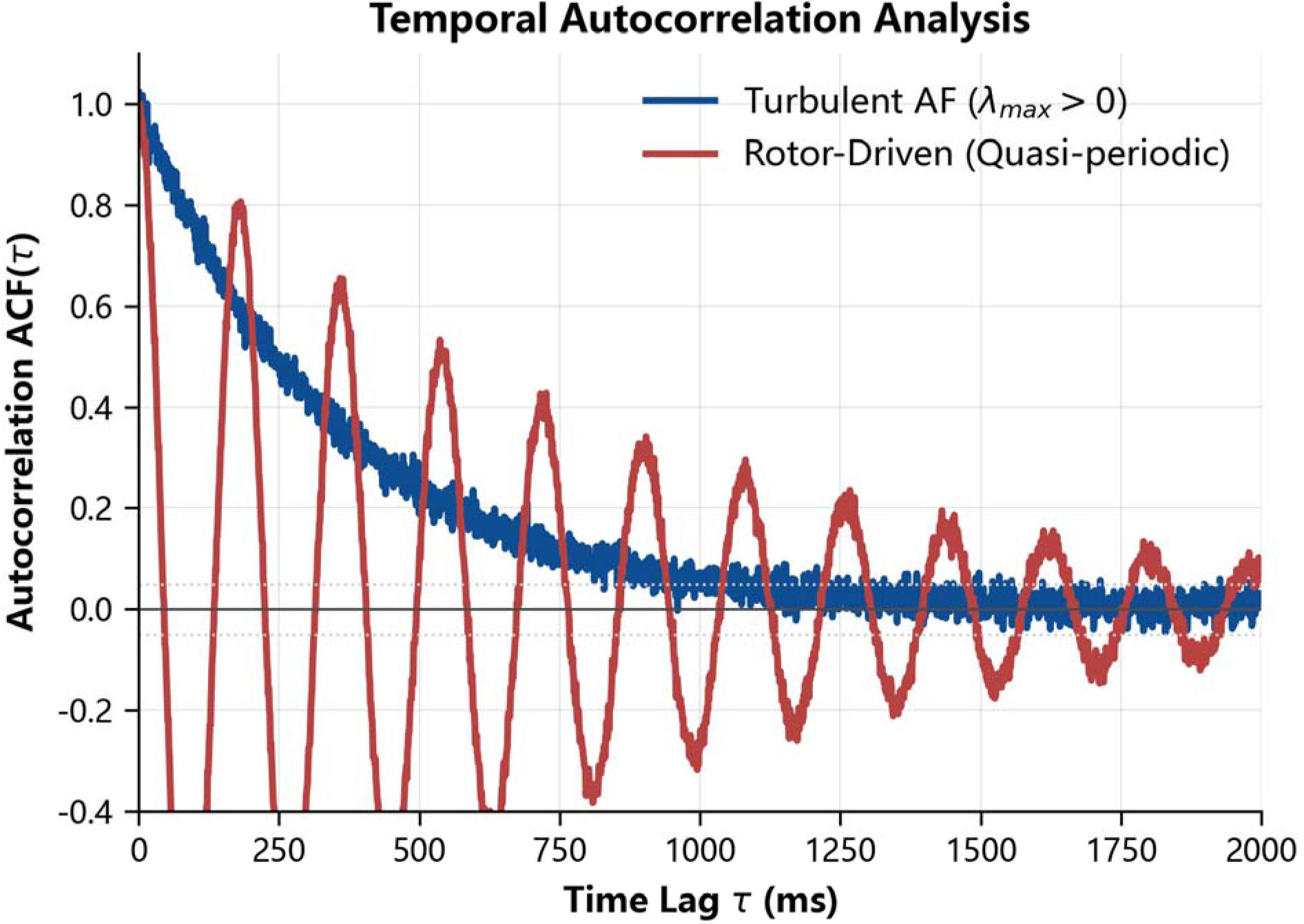
Autocorrelation function of the wavelet-count time series. The turbulence-like state showed rapid exponential decay, with a correlation time of approximately 350 ms, whereas the rotor-like state retained oscillatory periodic memory.

**Table 5.** Stage-wise statistics of raw wavelet-count time series.

| Stage | Mean wavelet count | Standard deviation | Minimum | Maximum | Interquartile range |
| --- | --- | --- | --- | --- | --- |
| Stage 0 | 0.45 | 0.84 | 0 | 5 | 0-1 |
| Stage 2 | 1.18 | 1.45 | 0 | 10 | 0-2 |
| Stage 4 | 4.94 | 1.91 | 2 | 8 | 3-6 |
| Stage 5 | 6.45 | 2.18 | 3 | 10 | 5-8 |

**Table 6.**
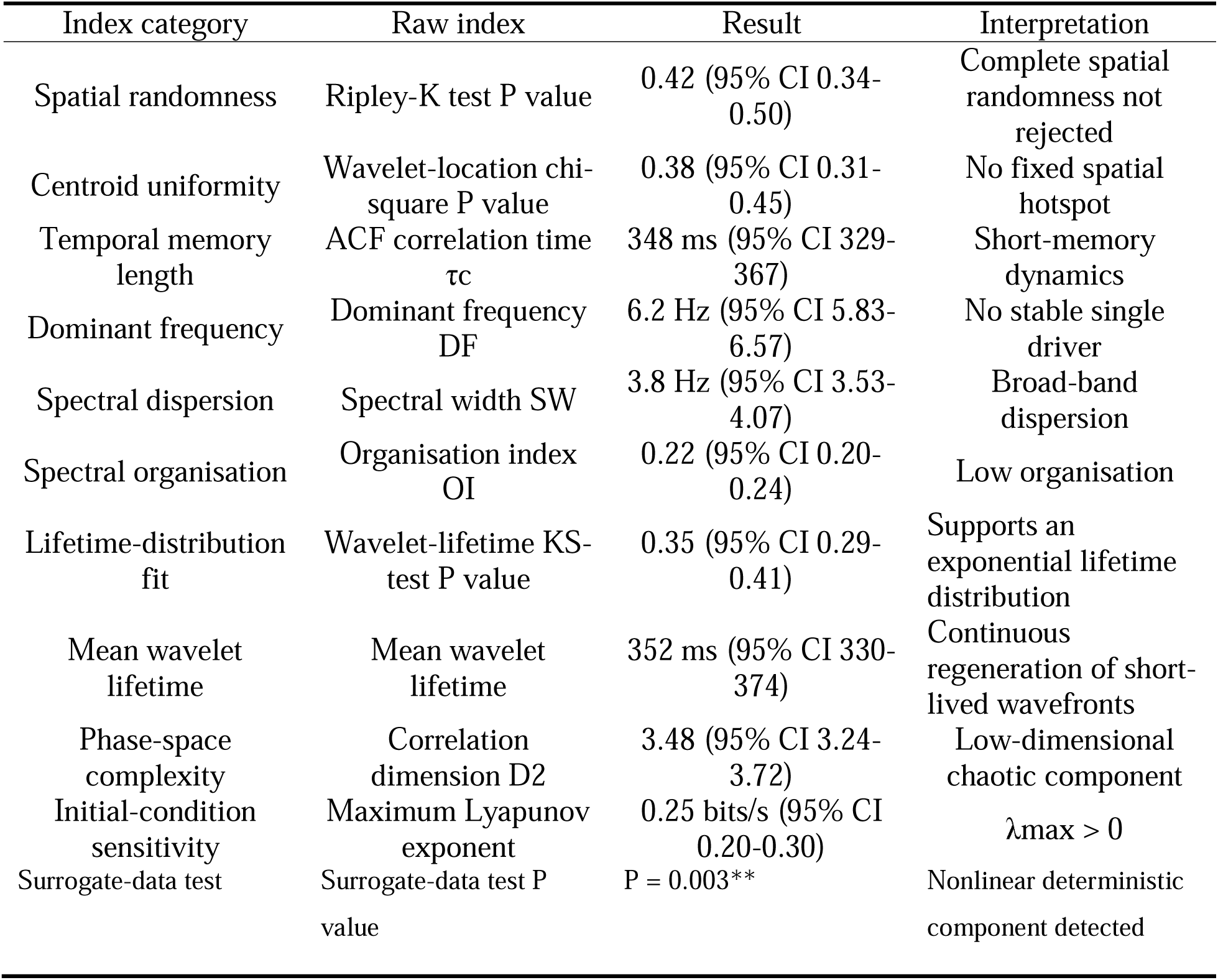
Spatial, temporal, spectral and chaotic indices of stage 5 turbulence-like electrical activity. P values were obtained from Ripley-K Monte Carlo envelope testing, wavelet-location chi-square testing, Kolmogorov-Smirnov testing of wavelet-lifetime distributions against the exponential distribution and surrogate-data testing, as indicated for each row. *P<0.05, **P<0.01 and ***P<0.001.

### Broad Spectrum and Positive Lyapunov Exponent Indicated Low-Organization Chaotic Components

The virtual-electrode wavelet spectrum showed that, in the turbulence-like state, energy was broadly distributed over 5-15 Hz without a stable single dominant frequency (Figure 10). Power-spectrum analysis of the wavelet-count time series showed broad gradual decay and no prominent dominant peak in the advanced turbulence-like state (Figure 11). Power-spectrum comparison across remodeling stages showed that stage 0 was dominated by a narrow-band ordered spectrum, stage 2 began to show spectral widening, and stages 4 and 5 gradually shifted to broad dispersed spectra (Figure 12). In stage 5, dominant frequency was 6.2 Hz, spectral width was 3.8 Hz and the organization index was only 0.22 (Table 6). Nonlinear-dynamical analysis showed a correlation dimension D2 of 3.48, maximum Lyapunov exponent of 0.25 bits/s and surrogate-data test P=0.003.

**Figure 10.**
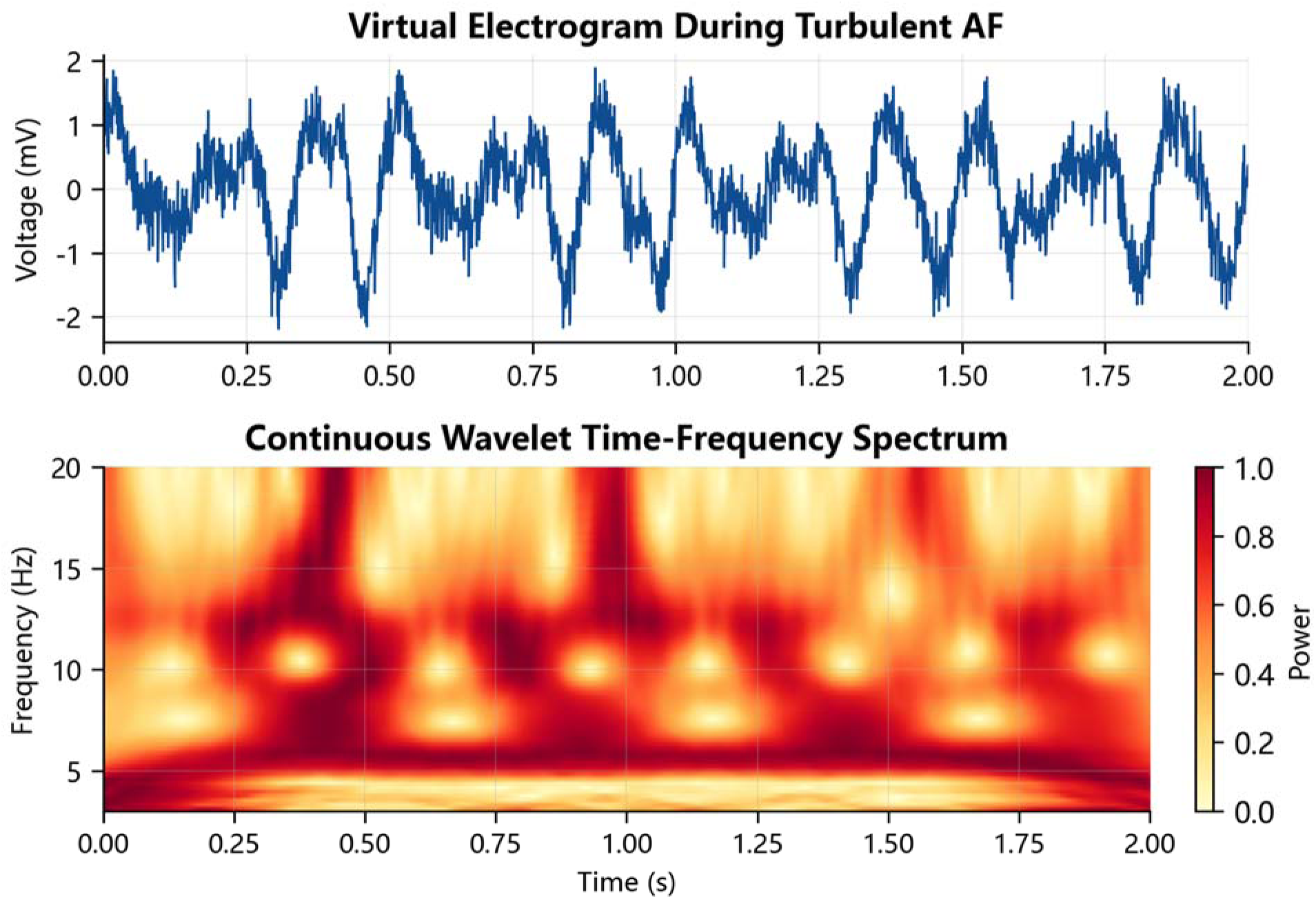
Virtual-electrode signal and continuous wavelet time-frequency spectrum. In the turbulence-like state, energy was broadly distributed across 5-15 Hz without a stable single dominant frequency.

**Figure 11.**
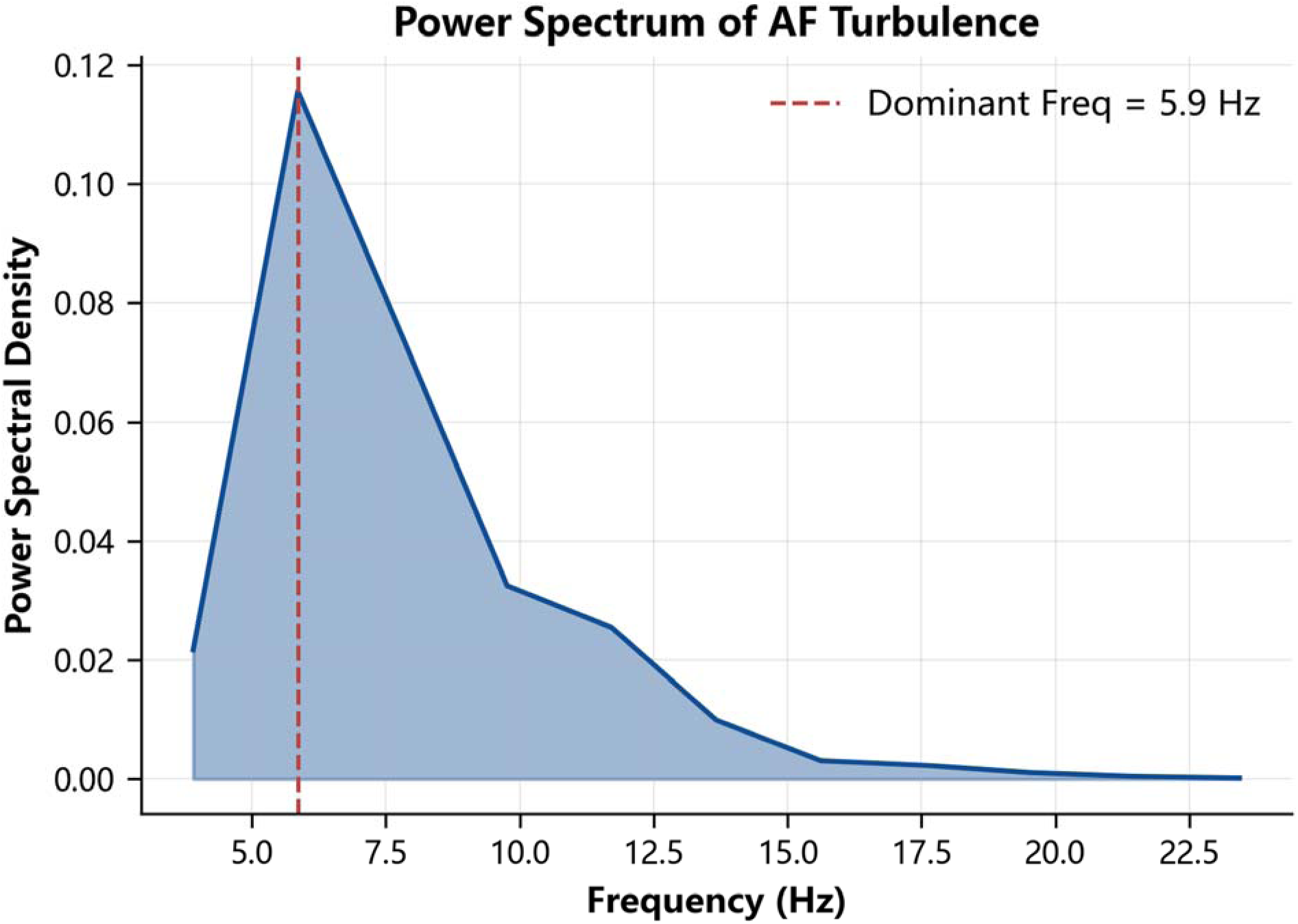
Power spectral density of the wavelet-count time series in the turbulence-like state. The spectrum showed broad gradual decay and lacked a prominent dominant peak, indicating dispersed frequency components.

**Figure 12.**
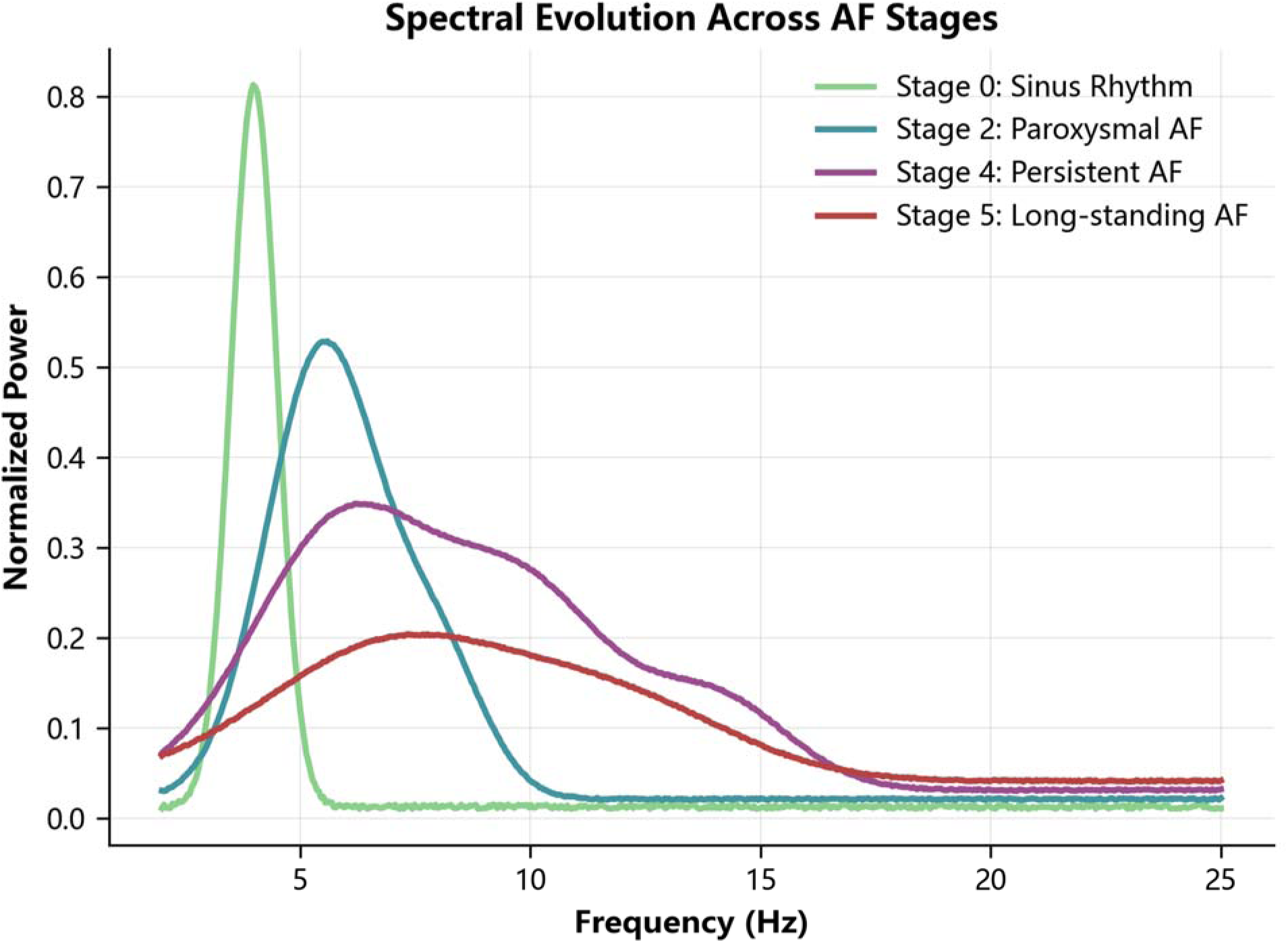
Power-spectrum comparison across remodelling stages. With increasing substrate remodelling, the spectrum evolved from a narrow-band single-peak pattern to a broad dispersed pattern.

### Virtual Ablation Showed That the Advanced State Did Not Depend on a Single Fixed Driver

Virtual ablation distinguished the intervention responses of rotor-like and turbulence-like states. In the stage 2 rotor-like state, targeted rotor ablation terminated 93.3% of episodes. In the stage 5 advanced substrate, random ablation terminated only 12.5%. When ablation area was 5%, persistence remained 87.5% in stage 5. Even when ablation area increased to 20%, persistence remained 62.3%. The post-ablation wavelet regeneration time constant was 1.15 s (Table 7).

**Table 7.** Virtual-ablation experiments and corresponding mechanistic states.

| Intervention<br>/mechanistic state | Endpoint | Result | Mechanistic implication |
| --- | --- | --- | --- |
| Stage 5 | Termination after random ablation | 12.5%(95% CI 10.4-14.6) | Decentralised state is insensitive to random local ablation |
| Stage 2 | Termination after rotor ablation | 93.3%(95% CI 91.8-94.8) | Local-driver state is sensitive to targeted ablation |
| Stage 5 | Wavelet-regeneration time constant | 1.15 s(95% CI 1.01-1.29) | Residual substrate rapidly regenerates wavelets after ablation |
| Stage 5 | Persistence after 5% ablation | 87.5%(95% CI 85.6-89.4) | Small-area ablation fails to terminate the advanced state |
| Stage 5 | Persistence after 20% ablation | 62.3%(95% CI 59.7-64.9) | Expanded ablation still did not fully alter the dynamical attractor |

### Multiscale Results Formed a Consistent Hypothesis-Testing Chain

Overall, stage 0 and stage 1 showed long wavelength, low wavelet capacity and low inducibility, corresponding to ordered or self-limited propagation. Stage 2 occupied a transition zone, in which wavebreak began to increase and rotor-like or local-driver behavior remained detectable. Stage 3 entered an early persistent-AF-like zone, in which Ncritical exceeded the threshold required for multiple wavelets and inducibility increased markedly. Stages 4 and 5 showed short wavelength, high wavelet capacity, spatial randomness, short-memory decay, spectral dispersion, positive nonlinear indices and poor response to random local ablation. These observations support the proposed sequence of substrate remodeling, wavelength shortening, wavebreak proliferation and decentralized multi-wavelet maintenance.

### Reverse Mechanistic Test by ERP Prolongation

ERP prolongation was used as a reverse mechanistic test of the critical-wavelength and wavelet-capacity chain. With fixed tissue area and conduction velocity, ERP prolongation increased effective wavelength according to lambda=ERP*CV and reduced Ncritical=A/lambda^2. Once lambda crossed the critical region near lambda50, wavelet capacity fell below the lower limit required for multi-wavelet maintenance and normalized maintenance risk declined sigmoidally (Figure 13). This result supports the directionality of the core mechanism: if wavelength shortening and increased wavelet capacity promote turbulence-like maintenance, then ERP prolongation, reduced repolarization dispersion or improved conduction safety should shift the system from a sustained-maintenance zone back toward a paroxysmal or terminating zone. This analysis remains mechanistic and requires future testing in true two– or three-dimensional reaction-diffusion models with large-scale S1-S2 coupling-interval scanning, and validation against drug therapy, upstream treatment or trigger-control clinical data.

**Figure 13.**
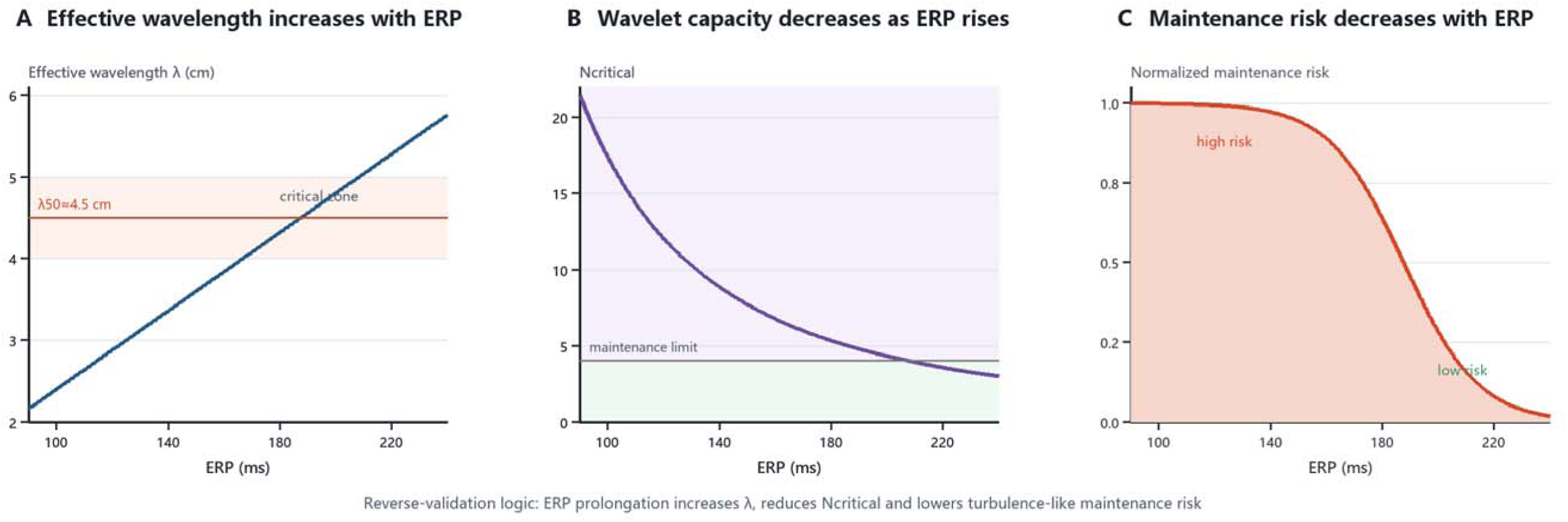
Reverse mechanistic test of ERP prolongation on critical wavelength, wavelet capacity and maintenance risk. (A) ERP prolongation increased effective wavelength and crossed the lambda50 threshold. (B) As lambda increased, Ncritical=A/lambda^2 decreased toward the lower limit for multi-wavelet maintenance. (C) Normalised maintenance risk declined sigmoidally with ERP prolongation.

## Discussion

### Computational Evaluation of the Turbulence-like Electrical Activity Hypothesis

This result supplies a model-derived dynamical transition marker rather than a clinical treatment cut-off: when the fixed 10 cm x 10 cm domain could accommodate multiple independent short-lived wavelets, the system became increasingly self-maintaining without a single fixed driver. The multi-wavelet theory proposed by Moe et al.9,10 provides the classical foundation for this phenomenon, while the present controlled model quantifies the relationship among wavelength, wavelet capacity and induction probability. The estimated lambda50 of approximately 4.5 cm is therefore a testable candidate for future substrate and ablation-stratification studies, including high-density mapping studies; it should not be used directly to select ablation targets or patients. The reduced-order design was chosen to expose the causal relation between effective wavelength and wavelet capacity, rather than to reproduce channel-specific kinetics or predict therapeutic effects.

Although this study did not systematically vary domain size, the consistency between lambda50 and Ncritical approximately equal to 5 suggests a capacity-based interpretation: the transition occurs when the tissue area can accommodate approximately five independent wavelets. If the transition occurs at a comparable Ncritical, a larger domain would be expected to require a proportionally longer threshold wavelength, scaling with the square root of A, whereas a smaller domain would require a shorter wavelength to reach the same Ncritical. This capacity-based view predicts that maintenance is governed by Ncritical rather than by absolute wavelength alone—a hypothesis that can be tested in future systematic area-scanning experiments.

The term turbulence-like in this study does not equate atrial tissue with a Navier-Stokes fluid. Cardiac tissue is an active excitable medium, in which propagation and recovery are sustained by ionic currents rather than by momentum-conserving flow. Near wavefront instability, reaction-diffusion systems can admit long-wave, phase-reduced descriptions of Kuramoto-Sivashinsky (KS)-type dynamics, in which nonlinear phase deformation competes with dissipative regularization to generate spatiotemporal irregularity.^38,39^ We therefore use the term turbulence-like as a dynamical descriptor of wavebreak, phase disorder and multi-wavelet renewal, not as a claim of hydrodynamic turbulence.

The reduced two-variable model was selected to isolate this mechanism. By holding geometry and model complexity fixed, it reduces parameter confounding from anatomical detail and ion-channel kinetics and makes the relation among ERP, CV, wavelength and wavelet capacity identifiable. This is a mechanism-isolation strategy, not a claim that structural and ionic heterogeneity are unimportant: recent work shows that heterogeneous ionic remodeling promotes wavefront instability,^40^ and contemporary clinical synthesis emphasizes the interacting contributions of triggers and substrate in AF maintenance.^41^ These factors should be incorporated in subsequent patient-specific tests of the present hypothesis.

### Relationship to Rotor, Spectral-Target and Ablation Studies

The present results do not reject the role of rotors or focal drivers in some AF stages. Instead, they suggest that the explanatory value of such mechanisms may change with disease stage and substrate remodeling. The stage 2 rotor-like state was highly sensitive to targeted ablation, whereas stage 5 was insensitive to random local ablation, indicating that early and advanced AF may occupy different dynamical regimes. Studies by Davidenko et al.^13^ and Gray et al.^14^ on spiral waves and spatiotemporal organization during fibrillation suggest that stable spirals, drifting rotors, multi-wave re-entry and chaotic wavebreak can be viewed as a continuum in excitable media. The persistent-AF ablation trial by Verma et al.^18^ showed that additional linear ablation or complex fractionated electrogram ablation was not necessarily superior to pulmonary-vein isolation alone. This clinical observation can be interpreted through decentralized maintenance: if wavelet regeneration depends on the whole substrate rather than on a single hotspot, the marginal effect of local target ablation decreases. Recent work by Kumagai et al.^19^ and Franco et al.^20^ explored persistent-AF targets based on dominant frequency and spatiotemporal dispersion, respectively, suggesting that a single mapping index is unlikely to capture complex AF organization fully. Scaglione et al.^21^ further showed that dynamic rotational activity in persistent AF requires spatial-temporal information for target identification. Therapeutically, this study supports a staged interpretation. When substrate fragmentation remains limited, dominant-frequency, rotational-activity or local-driver mapping may have high therapeutic value. Once the substrate has crossed the critical-wavelength threshold and acquired stable multi-wavelet capacity, treatment may need to shift from finding one source to changing the global conditions that allow turbulence-like maintenance, including prolonging effective wavelength, reducing heterogeneity gradients, reducing wavebreak-generating boundaries and controlling risk factors that promote remodeling.

### A Unified Explanation for Physiological and Pathological AF

Within this framework, AF initiation and maintenance should be separated conceptually. Paroxysmal or self-limited AF can occur in a structurally near-normal atrium if a premature atrial beat happens to fall within the physiological vulnerable window created by heterogeneous repolarization recovery. At that moment, some tissue remains refractory, some conducts slowly and some has recovered excitability, making wavefronts prone to one-way block and slow detouring around refractory borders. Pulmonary-vein sleeves are important trigger sources for paroxysmal AF, and premature beats from the pulmonary veins or similar sites that cross the vulnerable window and trigger instability can be defined as perturbation sources or, clinically, culprit premature beats.22 Alcohol, hyperthyroidism, sympathetic activation, hypoxia, electrolyte disturbance and dehydration may widen the vulnerable window or increase trigger probability by shortening ERP, increasing repolarization dispersion, reducing conduction safety or increasing trigger frequency. Abstinence from alcohol, restoration of euthyroid state and correction of acute triggers may therefore move the system back below the vulnerability threshold.4,23,24

By contrast, persistent and long-standing persistent AF are more dependent on increased pathological substrate capacity. The continuous stage 0 to stage 5 model indicates that, as effective wavelength shortens and N_critical_ increases, the same perturbation no longer merely triggers a transient event, but is more readily amplified by the remodelled substrate into self-sustaining multi-wavelet dynamics. In other words, the perturbation source answers why AF starts at that moment, the physiological vulnerable window explains why initiation is possible, and substrate capacity explains why AF does not self-terminate. This framework may explain heterogeneity in clinical treatment response. In perturbation-source-dominant cases, pulmonary-vein isolation, culprit premature beat ablation and trigger control may be particularly relevant. In patients with substantially increased substrate capacity, eliminating triggers alone may be insufficient because residual refractory dispersion, fibrotic boundaries and slow-conduction pathways can support recurrence. These cases may require substrate modification, risk-factor management and more comprehensive rhythm-control strategies.

The counter-pacing experiment extends the perturbation-source, vulnerable-window and substrate-capacity framework into an interventional dimension. Clinically, a culprit premature atrial beat within the atrial vulnerable window may be understood as an atrial-vulnerable-window analogy to the R-on-T phenomenon, but it should not be equated with a P wave falling on the surface ECG T wave. This framework also emphasizes that the timing and morphology of the initiating premature beat should be analysed rather than treated as a generic trigger. Premature atrial activity occurring during vulnerable atrial recovery may have different clinical implications depending on its morphology and anatomical origin; for example, a pattern suggesting a superior vena cava source would support considering superior vena cava isolation in addition to conventional pulmonary-vein isolation in appropriately selected patients. If an implanted device or catheter system can detect a premature atrial beat with a specific prematurity index, spatial origin and morphology in real time, local low-energy counter-pacing delivered 20-35 ms after the culprit beat may prevent AF initiation through wavefront collision, local phase resetting and transient filling of the vulnerable window. This strategy is not a replacement for pulmonary-vein isolation or culprit premature beat ablation. Rather, it represents a complementary path between trigger prevention and substrate modification. Its clinical value will require validation in three-dimensional patient-specific models, ex vivo atrial tissue, in vivo animal experiments and prospective device studies.

### AF Turbulence Index and a Treatment-Stratification Framework

Translationally, critical wavelength, Ncritical, Ripley-K spatial randomness, ACF short memory, spectral width and virtual-ablation response could be integrated into an atrial fibrillation turbulence index (AFTI). When AFTI is low, episodes are more likely to be perturbation-source dominant, and treatment may focus on pulmonary-vein isolation, culprit premature beat ablation and control of triggers such as alcohol, hyperthyroidism and sympathetic activation. When AFTI is intermediate, both trigger and substrate are likely to contribute, and high-density mapping of key slow-conduction or dispersion regions may be combined with PVI, together with strategies that prolong ERP or reduce repolarization heterogeneity. When AFTI is high, substrate capacity and decentralized multi-wavelet maintenance are likely to dominate, and trigger elimination alone may be inadequate. Greater emphasis may be required on substrate modification, risk-factor management and upstream therapy. AFTI is currently a testable stratification hypothesis derived from computational results and should not replace clinical evidence, but it provides a quantitative framework for patient-specific digital atrial models and prospective validation.

### Implications for Personalised Modeling and Multimodal Translation

As a mechanistic computational study, the present work provides a transferable index system for future patient-specific models. Horrach et al.^25^ emphasized that fibrosis is not merely structural deposition, but generates arrhythmogenic substrate through changes in cellular coupling, ion channels and conduction pathways. Studies by Gharaviri et al.^26^ and Zanchi et al.^27^ suggest that high-density mapping and computational simulation can link conduction velocity, low-voltage regions and AF vulnerability. Buonocunto et al.^28^ highlighted that a key challenge for cardiac electrophysiology digital twins is translating real structure, boundary conditions and mechanistic parameters into models that predict treatment response. Yamamoto et al.^29^ and Kulathilaka et al.^30^ further showed that patient-specific computational platforms may help identify arrhythmogenic substrate from biatrial lesion evolution and micrometer-scale structure. Jaffery et al.^31^ explored automated ablation-lesion mask generation in virtual persistent-AF cohorts. Compared with these studies, the present work has the advantage of a controlled parameter space, which clearly shows the causal chain between wavelength threshold and multi-wavelet capacity. Its limitation is that it does not yet incorporate real three-dimensional anatomy, anisotropic fibre orientation or patient-specific fibrosis distribution.

AF is not only an electrical disorder, but is also accompanied by changes in left atrial flow, mechanics and metabolic environment. Vogl et al.^32^ showed that catheter ablation changes left atrial geometry and hemodynamics, but velocity, wall shear stress and vortex-pattern responses vary across individuals. Shen et al.^33^ linked left atrial turbulent shear stress, wall pressure and calcium-activated potassium-channel expression, suggesting that mechanical signals may affect electrophysiological remodeling. Sekine et al.^34^ showed that four-dimensional flow magnetic resonance imaging can identify non-physiological left atrial flow using velocity, stasis and vortex quantification. Computational fluid dynamics studies by Parker et al.^35^ and Corti et al.^36^ suggest quantifiable relationships among flow stasis, vortex structure and thrombosis risk. Lin et al.^37^ found that successful AF ablation improves but does not fully reverse abnormalities in left atrial mechanics and energy loss. Future studies that map λ50, Ripley-K, ACF, spectral width and virtual-ablation response onto real patients, and validate them against high-density mapping, late-gadolinium-enhancement MRI, four-dimensional flow MRI and real ablation outcomes, may help distinguish AF states that remain modifiable by local targets from those already maintained by global substrate capacity.

### Limitations and Outlook

This study has several limitations. The model used an idealized two-dimensional, isotropic 10 cm x 10 cm tissue sheet and did not incorporate three-dimensional atrial anatomy, wall thickness, anisotropic conduction, pulmonary-vein sleeves, endocardial-epicardial dissociation or patient-specific boundaries. The two-variable FitzHugh-Nagumo system was used to isolate wavelength-dependent dynamics; it does not reproduce Na+-channel gating, CV/ERP restitution, calcium handling, autonomic regulation, metabolic remodeling or electromechanical coupling. Fibrosis was represented by a randomized spatial mask rather than replacement, infiltrative, endomysial, patchy or confluent fibrosis, and the present study did not compare how these spatial patterns alter the transition. The parameter ranges were literature informed but not patient specifically calibrated, and no experimental or clinical mapping dataset was used for external validation. Virtual ablation did not represent catheter energy delivery, lesion continuity, tissue oedema, conduction recovery or postprocedural inflammation. Future work should test non-uniform fibrosis patterns and determine whether frequency loading alone (S1-S1, without an S2 premature beat) can cross the model-derived transition, together with more detailed ionic and three-dimensional models validated against high-density mapping and patient-specific models.

A critical methodological consideration is that the absolute transition near λ50=4.5 cm was derived from the standardized 10 cm × 10 cm (100 cm²) simulation domain and is therefore domain-size dependent. The area-adjusted quantity Ncritical = A/λ² provides the appropriate mechanistic reference across domain sizes, whereas the 4.5 cm value should be interpreted only as a reference for the standard domain. If the transition occurs at a comparable Ncritical and a patient-specific effective excitable atrial area can be estimated, a first-order scaling hypothesis would be λcrit(patient) ≈ 4.5 cm × √(Apatient/100 cm²), where Apatient is expressed in cm²; for example, 150 cm² would yield approximately 5.5 cm under this unvalidated scaling. This relationship is a hypothesis for patient-specific modeling and mapping, not a validated clinical correction, because left-atrial surface area is only a proxy for effective excitable tissue and three-dimensional boundaries, anisotropy and fibrosis geometry may alter the relationship.

### Conclusions

Within a controlled two-dimensional excitable-medium model, progressive shortening of effective wavelength was associated with increased wavelet capacity, inducibility and multi-wavelet maintenance. The model-derived lambda50 of approximately 4.5 cm is a hypothesis-generating transition marker for future experimental, mapping and patient-specific modeling studies, not a current clinical treatment threshold.

## Acknowledgments

The authors thank the clinical electrophysiology and cardiovascular imaging teams for their support in discussing the translational framing of this simulation study.

## Author Contributions

X.C. conceived the study, built the computational workflow, analyzed the simulation outputs and drafted the manuscript. D.L. supervised the study and served as co-corresponding author. Q.Q., J.X., X.W., M.L., C.J., R.T., T.L., X.Z., H.Y., Z.X., K.H., P.G. and B.F. contributed to interpretation, clinical framing and manuscript revision. All authors reviewed and approved the final version.

## Disclosures

The authors have reported that they have no relationships relevant to the contents of this paper to disclose.

## Sources of Funding

This work was supported by the National Science Foundation of China (82151306, 82070339, 82400378), the Noncommunicable Chronic Diseases-National Science, Technology Major Project (2023ZD0504200, 2023ZD0513800), and the Beijing Physician Scientist Training Project (BJPSTP-2025).

## Data Availability

The unified original simulation data and R statistical analysis code supporting the quantitative results, tables and non-schematic figures are provided with this submission as source data files.

## Ethics Statement

Institutional review board or ethics committee approval was not required because this study used only computational simulations and did not involve human participants, animals, identifiable personal data, or biological specimens.

## Use of Artificial Intelligence

GPT-5.5 was used to assist with figure-design conceptualization and English-language polishing. It was not used to generate simulation data, perform statistical analyses or interpret results. The authors reviewed, edited and approved all AI-assisted outputs and take full responsibility for the accuracy, integrity and final content of the manuscript.

## Nonstandard Abbreviations and Acronyms

ACF: autocorrelation function
AF: atrial fibrillation
AFTI: atrial fibrillation turbulence index
APB: atrial premature beat
CI: coupling interval
CPBI: culprit premature beat index
CV: conduction velocity
ERP: effective refractory period
VWI: vulnerable-window index

## Supplementary Material

**Supplementary Table S1.**
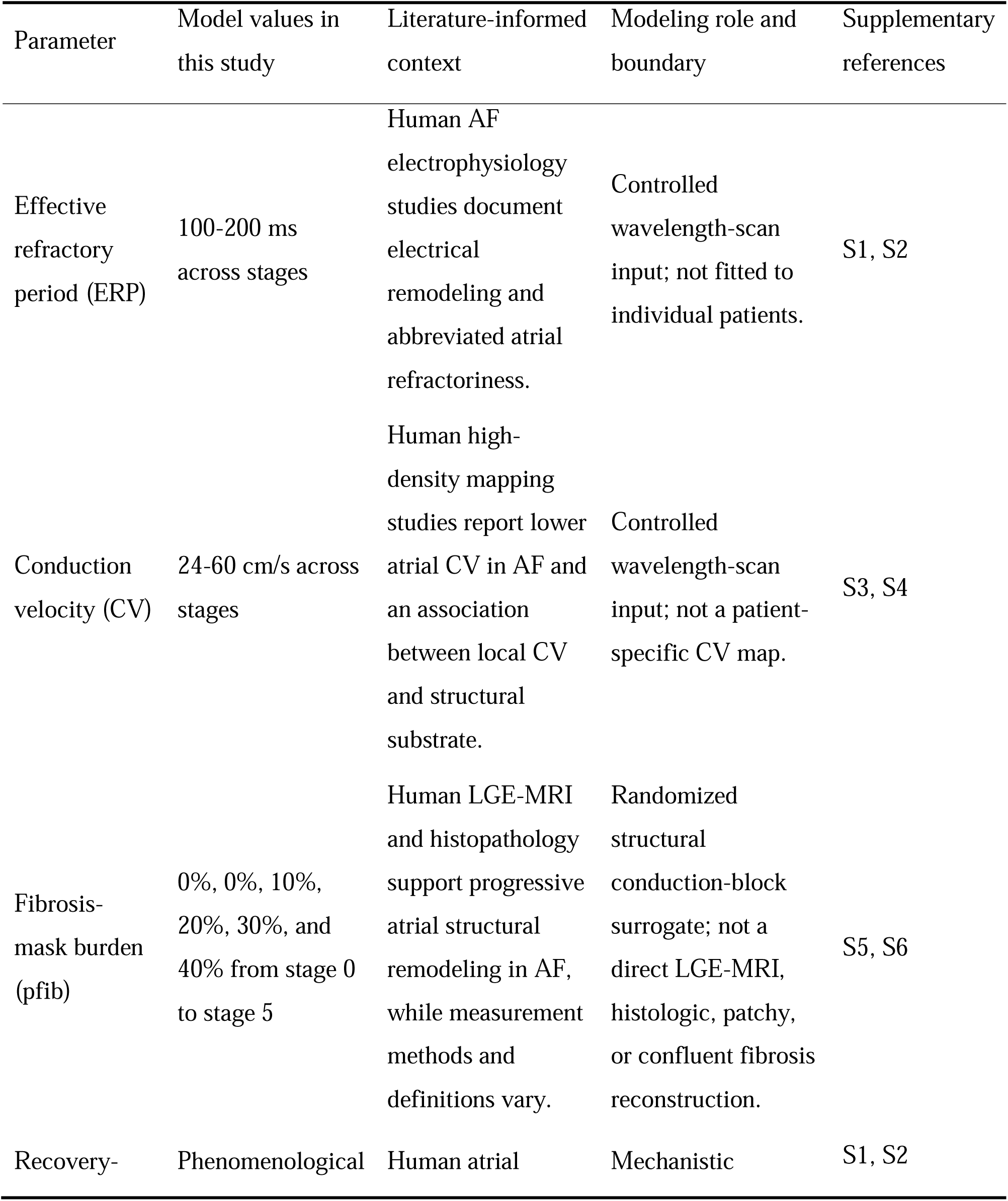
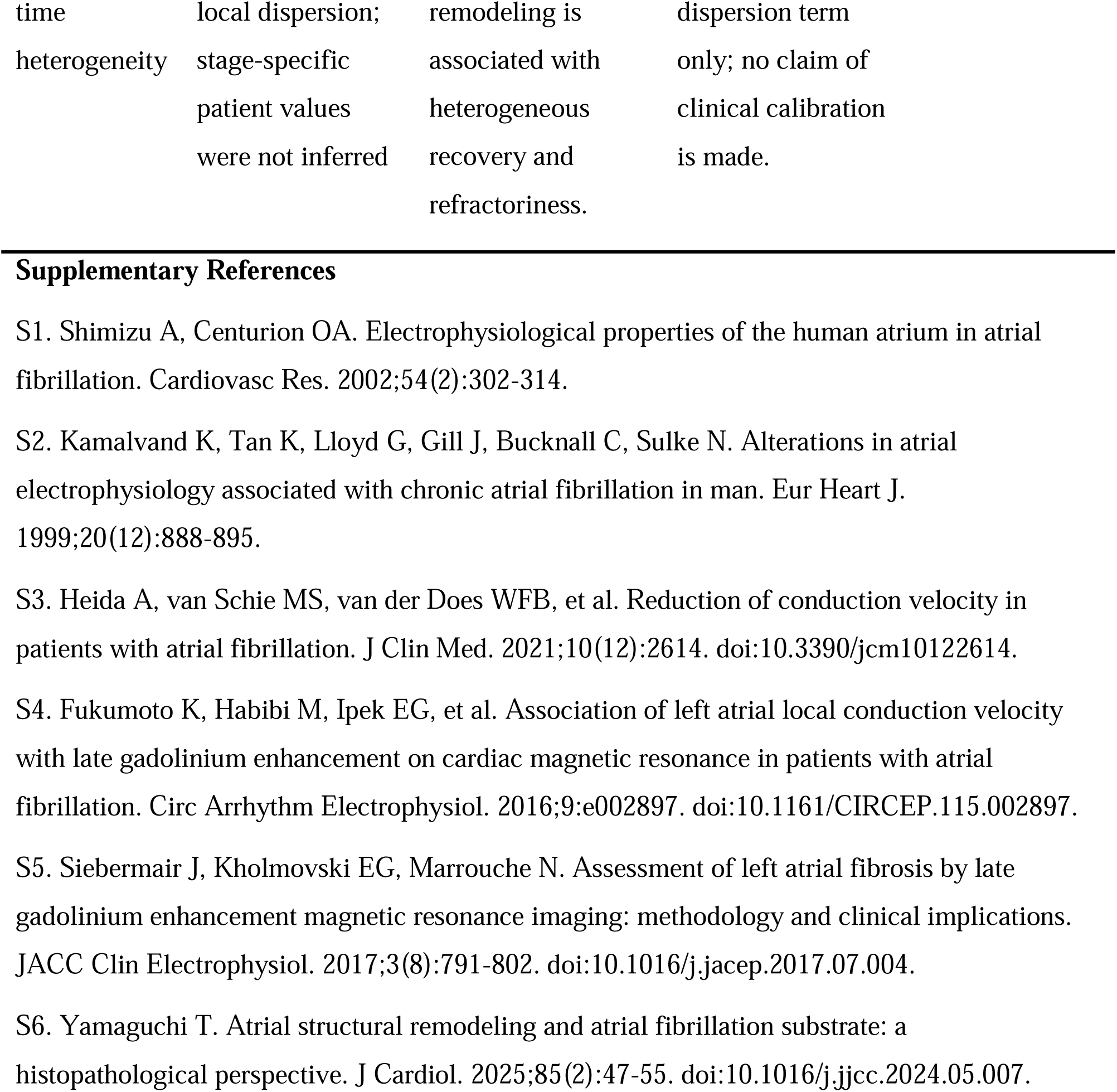
Clinically informed, literature-based substrate parameter context and modeling role.

| Parameter | Model values in this study | Literature-informed context | Modeling role and boundary | Supplementary references |
| --- | --- | --- | --- | --- |
| Effective refractory period (ERP) | 100-200 ms across stages | Human AF electrophysiology studies document electrical remodeling and abbreviated atrial refractoriness. | Controlled wavelength-scan input; not fitted to individual patients. | S1, S2 |
| Conduction velocity (CV) | 24-60 cm/s across stages | Human high-density mapping studies report lower atrial CV in AF and an association between local CV and structural substrate. | Controlled wavelength-scan input; not a patient-specific CV map. | S3, S4 |
| Fibrosis-mask burden (pfib) | 0%, 0%, 10%, 20%, 30%, and 40% from stage 0 to stage 5 | Human LGE-MRI and histopathology support progressive atrial structural remodeling in AF, while measurement methods and definitions vary. | Randomized structural conduction-block surrogate; not a direct LGE-MRI, histologic, patchy, or confluent fibrosis reconstruction. | S5, S6 |
| Recovery- | Phenomenological | Human atrial | Mechanistic | S1, S2 |

|  |  |  |  |
| --- | --- | --- | --- |
| time | local dispersion; | remodeling is | dispersion term |
| heterogeneity | stage-specific | associated with | only; no claim of |
|  | patient values | heterogeneous | clinical calibration |
|  | were not inferred | recovery and | is made. |
|  |  | refractoriness. |  |

## Notes

### Competing Interest Statement

The authors have declared no competing interest.

### Summary of Updates

The manuscript has been revised solely to remove the suffix from the first author's name. No other changes have been made.

## References

1. Lippi G, Sanchis-Gomar F, Cervellin G. Global epidemiology of atrial fibrillation: An increasing epidemic and public health challenge. Int J Stroke. 2021;16(2):217–221. doi: 10.1177/1747493019897870.

2. Shi S, Tang Y, Zhao Q, et al. Prevalence and risk of atrial fibrillation in China: A national cross-sectional epidemiological study. Lancet Reg Health West Pac. 2022;23:100439. doi: 10.1016/j.lanwpc.2022.100439.

3. Lane DA, Andrade JG, Arbelo E, et al. Atrial fibrillation. Lancet. 2026;407(10532):1000-1013. doi: 10.1016/S0140-6736(25)02166-X.

4. Joglar JA, Chung MK, Armbruster AL, et al. 2023 ACC/AHA/ACCP/HRS Guideline for the Diagnosis and Management of Atrial Fibrillation. Circulation. 2024;149(1):e1–e156. doi: 10.1161/CIR.0000000000001193.

5. Van Gelder IC, Rienstra M, Bunting KV, et al. 2024 ESC Guidelines for the management of atrial fibrillation developed in collaboration with the European Association for Cardio-Thoracic Surgery. Eur Heart J. 2024;45(36):3314–3414. doi: 10.1093/eurheartj/ehae176.

6. Chinese Medical Association Cardiovascular Diseases Branch CBESHRB. Guidelines for the Diagnosis and Treatment of Atrial Fibrillation in China. Chinese Journal of Cardiology, 2023.

7. Brown SM, Larsen NK, Thankam FG, et al. Regulatory role of cardiomyocyte metabolism via AMPK activation in modulating atrial structural, contractile, and electrical properties following atrial fibrillation. Can J Physiol Pharmacol. 2021;99(1):36–41. doi: 10.1139/cjpp-2020-0313.

8. Mines GR. On dynamic equilibrium in the heart. J Physiol. 1913;46(4-5):349–383. doi: 10.1113/jphysiol.1913.sp001596.

9. Moe GK, Abildskov JA. Atrial fibrillation as a self-sustaining arrhythmia independent of focal discharge. Am Heart J. 1959;58(1):59–70. doi: 10.1016/0002-8703(59)90274-1.

10. Moe GK, Rheinboldt WC, Abildskov JA. A COMPUTER MODEL OF ATRIAL FIBRILLATION. Am Heart J. 1964;67:200–220. doi: 10.1016/0002-8703(64)90371-0.

11. Cox JL, Canavan TE, Schuessler RB, et al. The surgical treatment of atrial fibrillation. II. Intraoperative electrophysiologic mapping and description of the electrophysiologic basis of atrial flutter and atrial fibrillation. J Thorac Cardiovasc Surg. 1991;101(3):406–426.

12. Wang Z, Pag P, Nattel S. Mechanism of flecainide’s antiarrhythmic action in experimental atrial fibrillation. Circ Res. 1992;71(2):271–287. doi: 10.1161/01.res.71.2.271.

13. Davidenko JM, Pertsov AV, Salomonsz R, et al. Stationary and drifting spiral waves of excitation in isolated cardiac muscle. Nature. 1992;355(6358):349–351. doi: 10.1038/355349a0.

14. Gray RA, Pertsov AM, Jalife J. Spatial and temporal organization during cardiac fibrillation. Nature. 1998;392(6671):75–78. doi: 10.1038/32164.

15. Chu X, Jiang X, Qiao Q, et al. Understanding Atrial Fibrillation Complexity Through the Lens of Turbulence Dynamics: Implications for Treatment Strategies. J Cardiovasc Electrophysiol. 2026;37(2):400–412. doi:10.1111/jce.70229.

16. FitzHugh R. Impulses and physiological states in theoretical models of nerve membrane. Biophys J. 1961;1(6):445–466. doi: 10.1016/S0006-3495(61)86902-6.

17. Nagumo J, Arimoto S, Yoshizawa S. An active pulse transmission line simulating nerve axon. Proc IRE. 1962;50(10):2061–2070. doi: 10.1109/JRPROC.1962.288235.

18. Verma A, Jiang CY, Betts TR, et al. Approaches to catheter ablation for persistent atrial fibrillation. N Engl J Med. 2015;372(19):1812–1822. doi: 10.1056/NEJMoa1408288.

19. Kumagai K, Hasegawa T, Kutsuzawa D, et al. Prospective Trial of Radiofrequency Catheter Ablation of High Dominant Frequencies After Pulmonary Vein Isolation in Non-Paroxysmal Atrial Fibrillation (PAD-AF Trial): A Multicenter, Randomized Clinical Trial. J Cardiovasc Electrophysiol. 2026. doi: 10.1111/jce.70357.

20. Franco E, Lozano-Granero C, Antonana-Ugalde S, et al. Ablation of visually identified spatiotemporal dispersion plus pulmonary vein isolation in persistent atrial fibrillation. Eur Heart J Open. 2026;6(2):oeag063. doi: 10.1093/ehjopen/oeag063.

21. Scaglione M, Geuna F, Spinoni EG, et al. A multicentre study on persistent atrial fibrillation ablation targeting rotational activity on top of pulmonary vein isolation: insights from an extensive mapping technique. Front Cardiovasc Med. 2026;13:1746001. doi: 10.3389/fcvm.2026.1746001.

22. Haissaguerre M, Jais P, Shah DC, et al. Spontaneous initiation of atrial fibrillation by ectopic beats originating in the pulmonary veins. N Engl J Med. 1998;339(10):659–666. doi: 10.1056/NEJM199809033391003.

23. Voskoboinik A, Kalman JM, De Silva A, et al. Alcohol Abstinence in Drinkers with Atrial Fibrillation. N Engl J Med. 2020;382(1):20–28. doi: 10.1056/NEJMoa1817591.

24. Reddy V, Taha W, Kundumadam S, Khan M. Atrial fibrillation and hyperthyroidism: A literature review. Indian Heart J. 2017;69(4):545–550. doi: 10.1016/j.ihj.2017.07.004.

25. Horrach CV, Bevis L, Nwanna C, et al. Atrial fibrosis in atrial fibrillation: Mechanisms, mapping techniques and clinical applications. J Physiol. 2025. doi: 10.1113/JP288680.

26. Gharaviri A, Vigneswaran V, Vickneson K, et al. Performance of atrial conduction velocity algorithms with error-prone clinical measurements for the identification of atrial fibrosis. Comput Biol Med. 2025;191:110119. doi: 10.1016/j.compbiomed.2025.110119.

27. Zanchi B, Gharaviri A, Bergonti M, et al. High-density atrial mapping, P-wave analysis, and computational simulations in Brugada syndrome: Enhancing the understanding of atrial fibrillation. Heart Rhythm O2. 2025;6(10):1621–1631. doi: 10.1016/j.hroo.2025.06.027.

28. Buonocunto M, Jung A, Meier S, et al. Moving towards digital twins for precision cardiac electrophysiology: overcoming technical and clinical challenges. Expert Rev Cardiovasc Ther. 2026. doi: 10.1080/14779072.2026.2674735.

29. Yamamoto C, Sakata K, Ali SY, et al. Arrhythmogenic substrates in atrial fibrillation and the role of ablation lesions: a longitudinal biatrial digital twin study. Cardiovasc Res. 2026;122(4):480–491. doi: 10.1093/cvr/cvag016.

30. Kulathilaka A, Sharma R, Kennelly J, et al. Structural determinants of re-entrant drivers in atrial fibrillation: insights from digital twins derived from 3D micrometer-resolution imaging of human heart. J Physiol. 2025. doi: 10.1113/JP288625.

31. Jaffery OA, Lopez-Barrera CE, Rodero C, et al. Automated generation of ablation lesion masks: a unison of electro and optic flow mapping for persistent AF virtual cohorts. Europace. 2026;28(1). doi: 10.1093/europace/euaf290.

32. Vogl BJ, Shaer AE, Van Zyl M, et al. Effect of catheter ablation on the hemodynamics of the left atrium: Hemodynamics of ablation. J Interv Card Electrophysiol. 2022;65(1):83–96. doi: 10.1007/s10840-022-01191-3.

33. Shen P, Ferdous M, Wang X, et al. A Detailed Study to Discover the Trade between Left Atrial Blood Flow, Expression of Calcium-Activated Potassium Channels and Valvular Atrial Fibrillation. Cells. 2022;11(9):1383. doi: 10.3390/cells11091383.

34. Sekine T, Nakaza M, Matsumoto M, et al. 4D Flow MR Imaging of the Left Atrium: What is Non-physiological Blood Flow in the Cardiac System?. Magn Reson Med Sci. 2022;21(2):293–308. doi: 10.2463/mrms.rev.2021-0137.

35. Parker L, Bollache E, Soulez S, et al. A multi-modal computational fluid dynamics model of left atrial fibrillation haemodynamics validated with 4D flow MRI. Biomech Model Mechanobiol. 2025;24(1):139–152. doi: 10.1007/s10237-024-01901-y.

36. Corti M, Zingaro A, Dede’ L, et al. Impact of atrial fibrillation on left atrium haemodynamics: A computational fluid dynamics study. Comput Biol Med. 2022;150:106143. doi: 10.1016/j.compbiomed.2022.106143.

37. Lin M, Hao L, Cao Y, et al. Successful catheter ablation of atrial fibrillation improves but not reverses the abnormalities of left atrial mechanics and energy loss. Echocardiography. 2019;36(4):752–760. doi: 10.1111/echo.14304.

38. Kuramoto Y. Instability and turbulence of wavefronts in reaction-diffusion systems. Prog Theor Phys. 1980;63(6):1885–1903. doi:10.1143/PTP.63.1885.

39. van Baalen G. Phase turbulence in the complex Ginzburg-Landau equation via Kuramoto-Sivashinsky phase dynamics. Commun Math Phys. 2004;247(3):613–654. doi:10.1007/s00220-004-1073-z.

40. O’Loughlin L, Dharmaprani D, Ganesan A, Mitchell L. Spatially heterogeneous ionic remodelling promotes wavefront instability in simulated atrial tissue. Front Netw Physiol. 2026;6:1849952. doi:10.3389/fnetp.2026.1849952.

41. Ciaccio EJ, Hsia HH, Talke M, et al. Contemporary concepts in the onset and maintenance of atrial fibrillation: Mechanisms, substrates, and clinical implications. Heart Rhythm. 2026. doi:10.1016/j.hrthm.2026.07.017.

## Supplementary References

S1. Shimizu A, Centurion OA. Electrophysiological properties of the human atrium in atrial fibrillation. Cardiovasc Res. 2002;54(2):302–314.

S2. Kamalvand K, Tan K, Lloyd G, Gill J, Bucknall C, Sulke N. Alterations in atrial electrophysiology associated with chronic atrial fibrillation in man. Eur Heart J. 1999;20(12):888–895.

S3. Heida A, van Schie MS, van der Does WFB, et al. Reduction of conduction velocity in patients with atrial fibrillation. J Clin Med. 2021;10(12):2614. doi:10.3390/jcm10122614.

S4. Fukumoto K, Habibi M, Ipek EG, et al. Association of left atrial local conduction velocity with late gadolinium enhancement on cardiac magnetic resonance in patients with atrial fibrillation. Circ Arrhythm Electrophysiol. 2016;9:e002897. doi:10.1161/CIRCEP.115.002897.

S5. Siebermair J, Kholmovski EG, Marrouche N. Assessment of left atrial fibrosis by late gadolinium enhancement magnetic resonance imaging: methodology and clinical implications. JACC Clin Electrophysiol. 2017;3(8):791–802. doi:10.1016/j.jacep.2017.07.004.

S6. Yamaguchi T. Atrial structural remodeling and atrial fibrillation substrate: a histopathological perspective. J Cardiol. 2025;85(2):47–55. doi:10.1016/j.jjcc.2024.05.007.

